# Health spending and vaccination coverage in low-income countries

**DOI:** 10.1101/2020.11.10.20229245

**Authors:** Francisco Castillo-Zunino, Pinar Keskinocak, Dima Nazzal, Matthew C Freeman

## Abstract

**Background:** Routine childhood immunization is a cost-effective way to save lives and protect people from disease. Some low-income countries (LIC) have achieved remarkable success in childhood immunization, despite lower levels of gross national income or health spending compared to other countries. We investigated the impact of financing and health spending on vaccination coverage across LIC and lower-middle income countries (LMIC).

**Methods:** Among LIC, we identified countries with high-performing vaccination coverage (LIC+) and compared their economic and health spending trends with other LIC (LIC-) and LMIC. We used cross-country multi-year linear regressions with mixed-effects to test financial indicators over time. We conducted three different statistical tests to verify if financial trends of LIC+ were significantly different from LIC- and LMIC; p-values were calculated with an asymptotic χ^2^ test, a Kenward-Roger approximation for F tests, and a parametric bootstrap method.

**Findings:** During 2014–18, LIC+ had a mean vaccination coverage between 91–96% in routine vaccines, outperforming LIC- (67–80%) and LMIC (83–89%). During 2000–18, gross national income and development assistance for health (DAH) per capita were not significantly different between LIC+ and LIC- (p > 0·13, p > 0·65) while LIC+ had a significant lower total health spending per capita than LIC- (p < 0·0001). Government health spending per capita per year increased by US$0·42 for LIC+ and decreased by US$0·24 for LIC- (p < 0·0001). LIC+ had a significantly lower private health spending per capita than LIC- (p < 0·012).

**Interpretation:** LIC+ had a difference in vaccination coverage compared to LIC- and LMIC that could not be explained by economic development, total health spending, nor aggregated DAH. The vaccination coverage success of LIC+ was associated with higher government health spending and lower private health spending, with the support of DAH on vaccines.

## Introduction

Routine childhood immunization has been one of the most cost-effective public health interventions to save lives and protect people from disease^1^. Investments in childhood immunization were estimated to yield a net return 44 times greater than costs during 2011–20, considering the value of people living longer/healthier lives and not needing treatment for vaccine-preventable diseases^2^.

Global immunization has significantly improved in the past decades, but there is still progress to be made in increasing coverage^3^ and understanding the impact of spending. External funding supporting vaccination efforts had a positive effect in DTP3 coverage from 1995–2004, while the effect was not significant in nations that reached a coverage greater than 65%^4^. Gavi, the Vaccine Alliance, received US$ 7·1 billion from governments and private organizations to support immunization and health systems of lower income and lower-middle income countries (LIC and LMIC) during 2016–20^5^. The goal imposed by the Global Vaccine Action Plan 2011–20 was to reach a 90% national coverage by the end of the period^6^. Despite the major efforts of international organizations and governments to improve vaccination coverage worldwide, DTP3 coverage has remained relatively consistent between 2010 (84%) and 2018 (86%)^7^.

Among LIC, we identified a subgroup (referred to as LIC+) with high-performance vaccination coverage compared to other LIC (referred to as LIC-) and LMIC. We investigated the time-varying differences of financial factors, such as DAH and government spending on health, between LIC+, LIC-, and LMIC.

Organizations, such as the Institute for Health Metrics and Evaluation (IHME), WHO, and UNICEF, have analyzed several health financing indicators with different disaggregation levels, e.g., funding by source and health focus area^8,9,10^. These efforts enabled researchers to project health spending patterns, identify and track spending trends, and do multivariate analysis combined with other health outcomes^11,12^.

The objective of this study is to understand the impact of health financing indicators on vaccination coverage rates of LIC. We used linear mixed-effects models^13^ to statistically compare LIC+, LIC-, and LMIC, regarding income, health spending, and vaccine spending, per capita or per birth. To the best of our knowledge, this is the first cross-country study that statistically analyzes the differences among LIC in terms of health financing over time and vaccination coverage success.

## Methods

### Data sources & processing

A summary of data sources can be found in Supplemental Materials (Table 1).

**Table 1:** Country groups with ISO3 country codes.

| LIC+ |  |  |  | LIC- |  |  |  |  |  |
| --- | --- | --- | --- | --- | --- | --- | --- | --- | --- |
| Burundi | BDI | Rwanda | RWA | Afghanistan | AFG | Haiti | HTI | Sierra Leone | SLE |
| Burkina Faso | BFA | Togo | TGO | Benin | BEN | Madagascar | MDG | Syrian Arab Rep. | SYR |
| Malawi | MWI | Tajikistan | TJK | Congo, Dem. Rep. | COD | Mali | MLI | Chad | TCD |
| Nepal | NPL | Tanzania | TZA | Ethiopia | ETH | Mozambique | MOZ | Uganda | UGA |
|  |  |  |  | Guinea | GIN | Niger | NER | Yemen, Rep. | YEM |
| LMIC |  |  |  |  |  |  |  |  |  |
| Angola | AGO | Ghana | GHA | Cambodia | KHM | Pakistan | PAK | Tunisia | TUN |
| Bangladesh | BGD | Honduras | HND | Lao PDR | LAO | Philippines | PHL | Ukraine | UKR |
| Bolivia | BOL | Indonesia | IDN | Morocco | MAR | Papua New Guinea | PNG | Uzbekistan | UZB |
| Côte d'Ivoire | CIV | India | IND | Myanmar | MMR | Sudan | SDN | Vietnam | VNM |
| Cameroon | CMR | Kenya | KEN | Nigeria | NGA | Senegal | SEN | Zambia | ZMB |
| Congo, Rep. | COG | Kyrgyz Rep. | KGZ | Nicaragua | NIC | El Salvador | SLV | Zimbabwe | ZWE |
| Egypt, Arab Rep. | EGY |  |  |  |  |  |  |  |  |

We used the WHO and UNICEF estimates of national infant immunization coverage (WU1^14^) from years 2000–18. We considered the first and third doses of diphtheria-tetanus-pertussis vaccines (DTP1 and DTP3), first dose of measles-containing vaccine (MCV1), bacillus Calmette-Guérin vaccine (BCG), and third dose of polio vaccine (Pol3) – these vaccines were picked because they target diseases included in the Expanded Programme on Immunization since 1977^15^. These estimates are based on government reports that are supplemented by survey results from the published and gray literature, in addition to feedback from local experts^16^.

From the World Bank’s world development indicators (WB1^17^) we obtained countries’ population and live birth rate, and used them to calculate per birth values. We also used GNI and gross domestic product (GDP) per capita. GNI values are expressed in US$ using World Bank Atlas method and GDP values are expressed in current US$.

Global health spending estimates for 195 countries and territories were obtained from publicly available data (IHME1^8^). We used the total health spending data disaggregated into government, out-of-pocket, prepaid private, and DAH (expressed in constant 2018 US$). DAH are the financial resources for the improvement and maintenance of health, transferred from major health development agencies to LIC and LMIC. We calculated private health spending as the sum of out-of-pocket and prepaid private health spending.

DAH estimates from 1990–2018 were disaggregated by health focus areas (IHME2^9^). We used DAH spent on newborn and child health; more specifically spent on vaccines (expressed in constant 2018 US$). DAH on vaccines include funding for routine immunization, new vaccines introduction, and support for delivery components such as cold chain optimization, systems strengthening, and human resources. We removed values marked as duplicates by IHME and data from 2018 since they contained few preliminary estimates.

From the immunization financing indicators (WU2^10^), from the WHO-UNICEF joint reporting form, we used total spending and government health spending on routine immunization; more specifically spending on vaccines used in routine immunization (expressed in constant 2010 US$). The spending on routine immunization includes the spending on vaccines used for routine immunization, plus delivery services – it does not include spending on vaccines used for supplementary activities (included in DAH on vaccines estimates). We removed data from 2018 since it is self-reported by countries and has not been audited by WHO.

### Selecting & grouping countries

We studied countries that were either LIC or LMIC in 2018 according to the World Bank’s income categorization. GNI per capita was US$1025 or less for LIC (31 countries), and between US$1026 and US$3995 for LMIC (47 countries). We removed countries with no World Bank income category nor GNI per capita reported during 2000–18. To avoid over-representing smaller nations, we removed countries whose population had never reached 5 million people. These criteria resulted in the selection of 23 LIC and 31 LMIC for the analysis.

As indicators of vaccination coverage success, we used DTP1 and DTP3 coverage; these metrics are widely used as proxies of routine vaccine system performance, since they have always been part of the Expanded Programme on Immunization^15^. DTP1 coverage indicates the initial engagement with the vaccine system, DTP3 coverage signifies the completion of the initial routine immunization, and the difference between DTP1 and DTP3 gauges the dropout or retention within the vaccine system^18^.

We selected subgroup LIC+ among LIC as follows: a mean DTP3 coverage above 85% during 2014– 18, and a mean DTP1–DTP3 dropout rate below 5% during the same period. These criteria lead to three country groups, summarized in Table 1. We picked the threshold of 85% DTP3 coverage by relaxing the Global Vaccine Action Plan goal of 90%^6^ – this increased the number of candidate LIC+ from 7 to 10. We also enforced 5% or less DTP1–3 dropout rate to have a strict standard for a country to be categorized in a high performing group; we consider the dropout rate a measure for the robustness of the vaccine system – this decreased the sample from 10 to 8 countries. However, we obtained similar results when using an alternative grouping criteria; results and plots can be found in Supplemental Materials.

### Comparing groups of countries

We compared LIC+ with LIC- and LMIC to understand the relationship between income and health spending trends and the vaccination coverage success. We evaluated if LIC+ had a higher income per capita than LIC-, spent more money in healthcare, received more DAH in general, received more DAH specific on vaccines, or if LIC+ invested more into vaccination coverage per birth. LMIC was used as a benchmark for comparison (considering their higher levels of spending).

The data used in the models span the years 2000 to 2018 (subject to data availability per country, summarized in Table 1 in Supplemental Materials), aligned with the launch of the Millennium Development Goals^19^ and the creation of Gavi. We used mixed-effects models^13^ that enable regression analysis with correlated variables, in this case by considering the random-effects of countries, and are unbiased estimators when data is missing at random^20^ (Table 1 in Supplemental Materials shows the percentage of data points missing for each variable.). Fixed-effects were implemented for each country group, to enable the comparison of the differences of magnitude and the trends between LIC+, LIC- and LMIC.

For significance testing, we used three different approaches: an asymptotic χ^2^ test, a Kenward-Roger approximation for *F* tests for reduction of mean structure, and a parametric bootstrap method^21^. These tests compute p-values to determine any significant differences between the group trends of LIC+ versus LIC- or LMIC; i.e., the p-value does not refer to the significance of a specific parameter of the regression but to the significance of all parameters combined. More details regarding the modeling and significance testing can be found in the section Mixed-effects models in Supplemental Materials. Data were analyzed in R with library *lme4*^*22*^.

## Results

We conducted a multi-year comparison of trends across LIC+, LIC-, and LMIC for the following indicators by year: vaccination coverage; GNI per capita; total, government, and private health spending per capita; DAH on vaccines per birth; and routine immunization spending per birth. In the figures, the trends for each country in LIC+ are indicated by their ISO3 country code. Table 2 in Supplemental Materials summarizes the intercepts and slopes (yearly change rate) for the trends. Table 3 in Supplemental Materials shows the p-values of the country group comparisons to test if their indicator trends were significantly different.

**Table 2:** Mean and standard deviation (SD) of vaccination coverage (2014–18)

| Vaccine | Mean (SD) vaccination coverage % |  |  |  |  |  |
| --- | --- | --- | --- | --- | --- | --- |
|  | LIC+ |  | LIC- |  | LMIC |  |
| DTP1 | 95.8 | (2.7) | 79.8 | (12.1) | 88.7 | (11.9) |
| DTP3 | 92.8 | (3.9) | 69.0 | (15.2) | 83.4 | (15.9) |
| MCV1 | 91.2 | (5.6) | 67.3 | (12.6) | 84.2 | (13.9) |
| BCG | 94.0 | (6.3) | 79.8 | (10.7) | 88.9 | (11.9) |
| Pol3 | 91.5 | (5.9) | 69.2 | (13.8) | 84.2 | (14.1) |

### Vaccination coverage

Figure 1 shows DTP3 coverage over time for each country in LIC+ and the aggregated trends (obtained by mixed-effect models) of country groups LIC- and LMIC. During 2014–18, LIC+ had a mean DTP3 coverage of 93%; surpassing LIC- (69%) and LMIC (83%). Table 2 shows the coverage (mean and standard deviation) summary for other mandatory vaccines proposed by WHO^15^. Every country in LIC+ outperformed the mean coverage of LIC- and LMIC for each studied mandatory vaccine.

**Figure 1:**
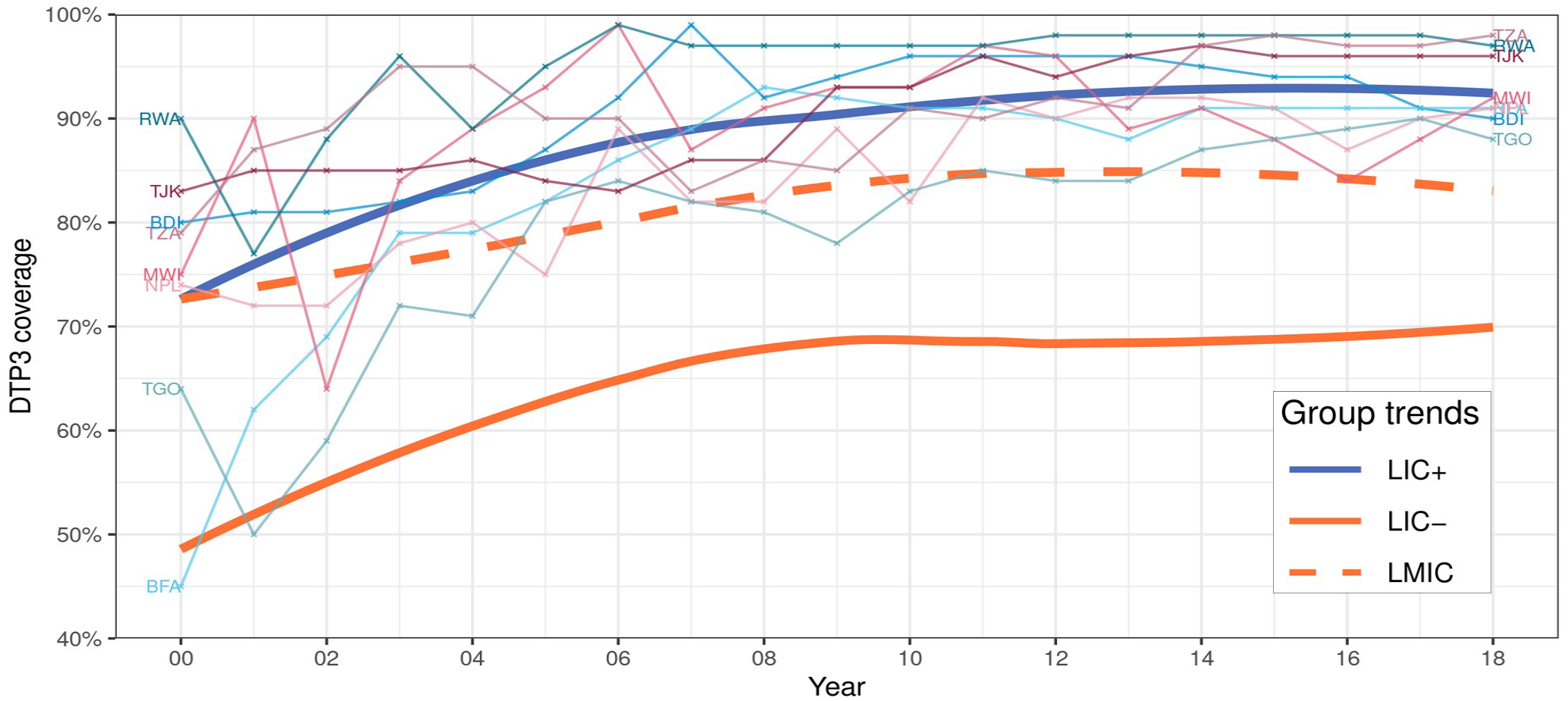
*DTP3* coverage of country groups. Data source: WU1. The trends of LIC+, LIC-, and LMIC were fitted by a local polynomial regression (a locally estimated scatter-plot smoothing; usually referred to as loess^23^). LIC+ countries (ISO3): Burundi (BDI), Burkina Faso (BFA), Malawi (MWI), Nepal (NPL), Rwanda (RWA), Togo (TGO), Tajikistan (TJK), and Tanzania (TZA).

### Income levels

Figure 2 shows the GNI per capita by year for each country in LIC+ and the aggregated trends (obtained by mixed-effect models) of country groups LIC- and LMIC. During 2000–18, the GNI and GDP per capita of LIC+ were significantly lower compared to LMIC (p < 0·0001, p < 0·0001), and also lower compared to LIC-although not significantly (p > 0·13, p > 0·42). The GNI per capita of LIC+ increased from US$199 to US$785, remaining consistently and significantly lower than that of LMIC (which increased from US$571 to US$2487), and slightly below LIC- (which increased from US$320 to US$831).

**Figure 2:**
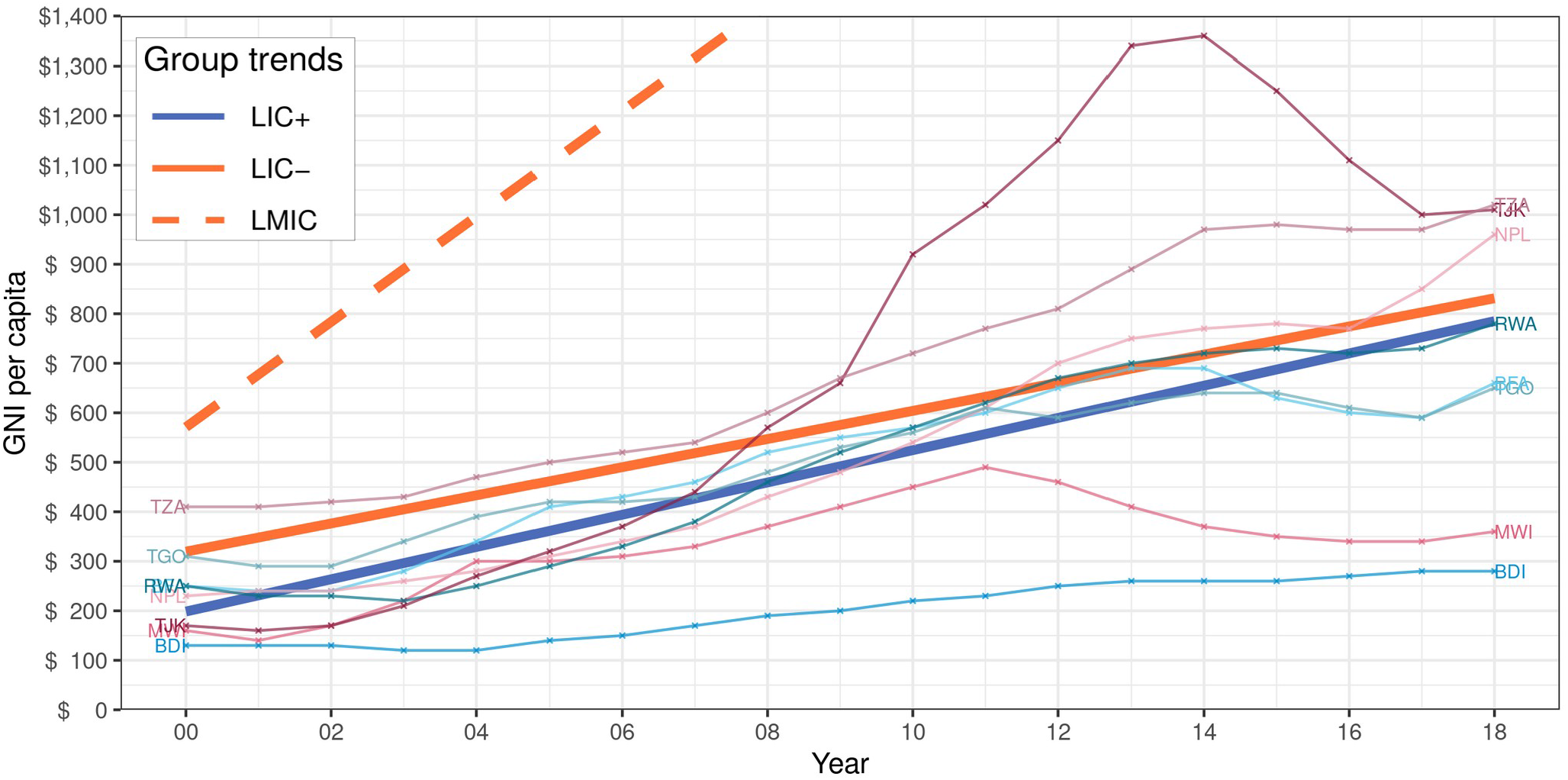
GNI per capita of country groups. Data source: WB1. The trends of LIC+, LIC-, and LMIC were fitted by linear mixed-effects models; note part of the LMIC trend was cut off for visibility. LIC+ countries (ISO3): Burundi (BDI), Burkina Faso (BFA), Malawi (MWI), Nepal (NPL), Rwanda (RWA), Togo (TGO), Tajikistan (TJK), and Tanzania (TZA).

### Health spending

Figure 3 shows the total, government, and private health spending per capita trends for country groups LIC- and LMIC (obtained by the mixed-effects models) and for each country in LIC+ during 2000–16.

**Figure 3:**
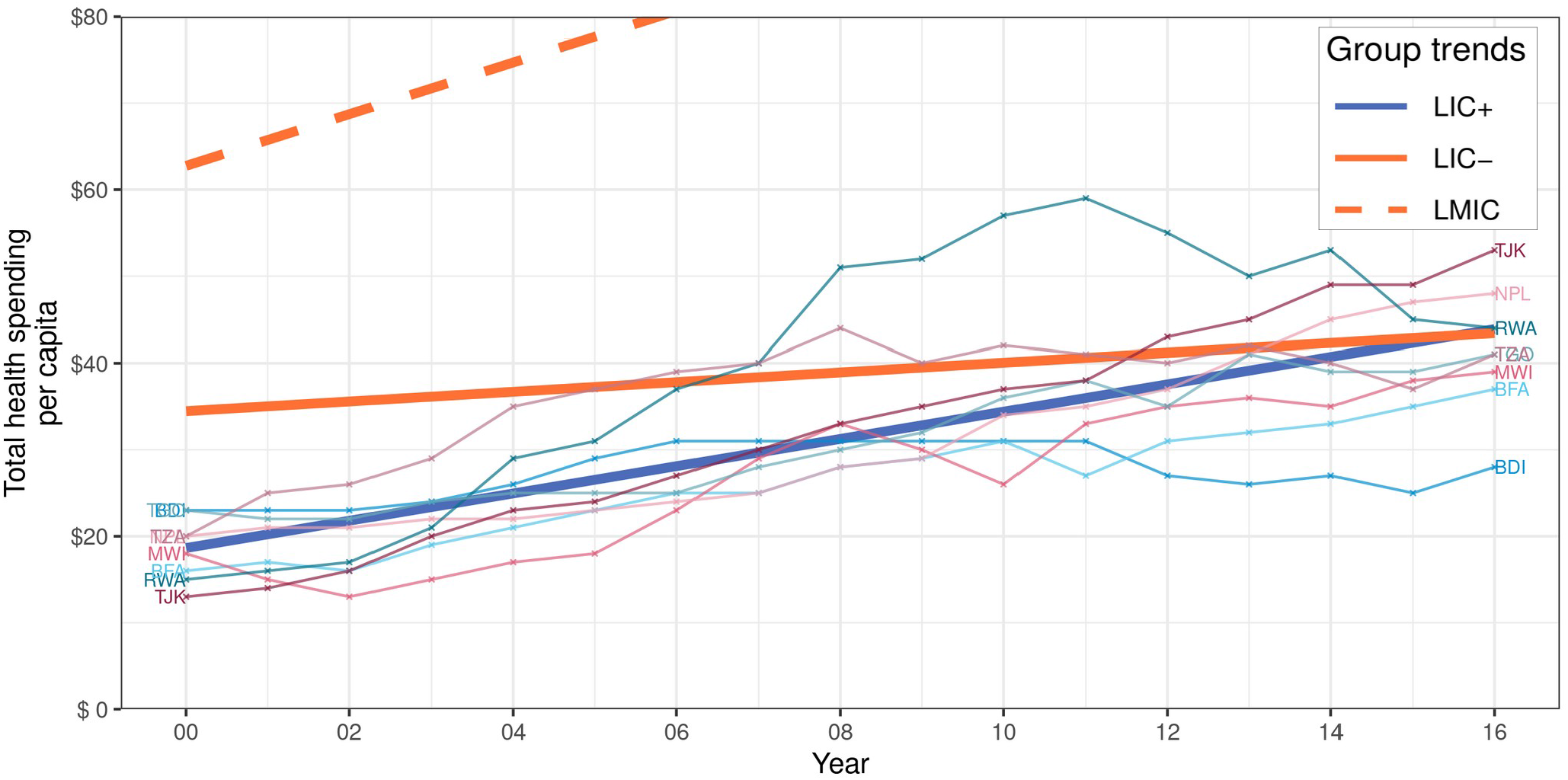
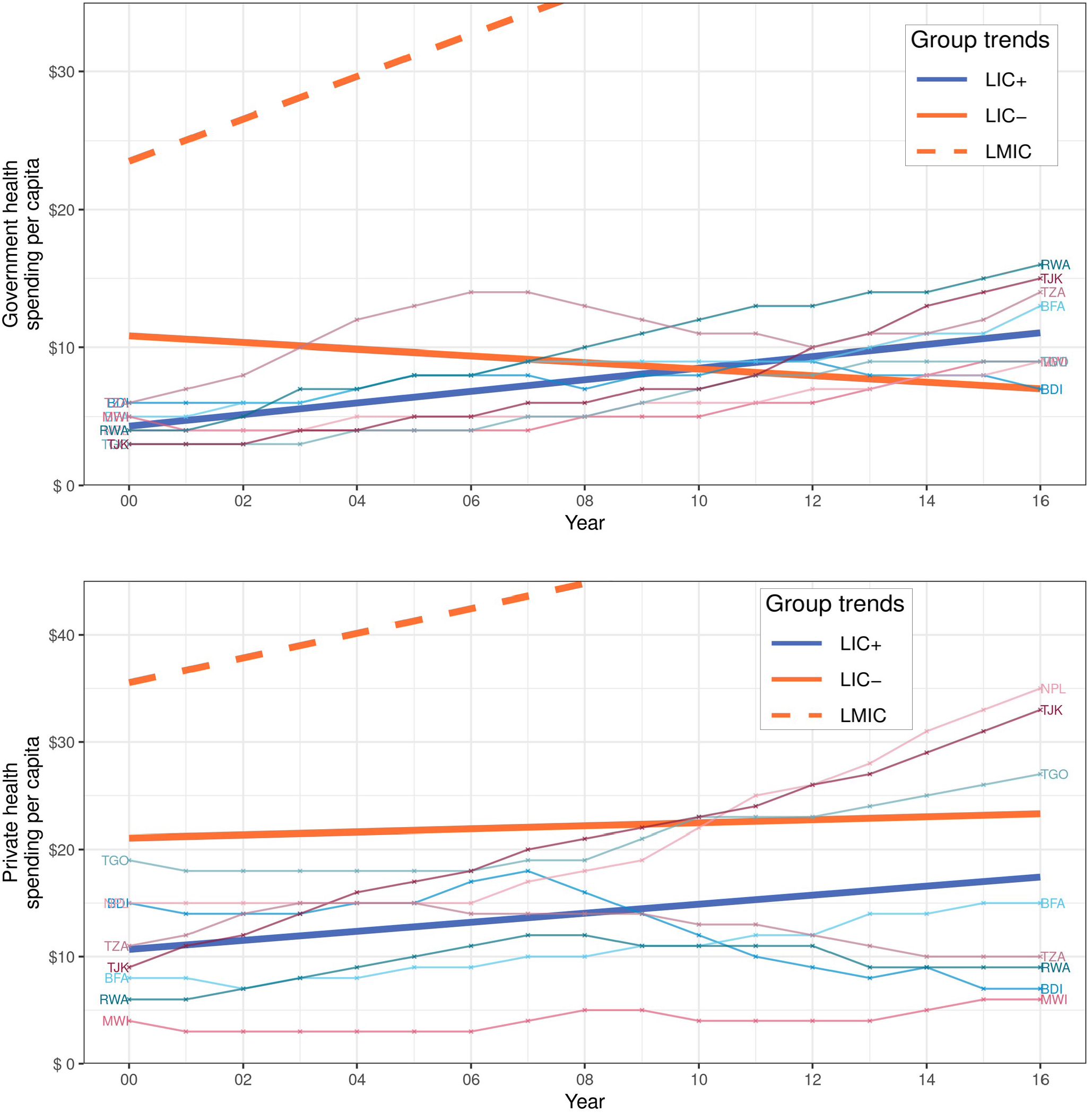
Total, government, and private health spending per capita of country groups. Data source: IHME1. The trends of LIC+, LIC-, and LMIC were fitted by linear mixed-effects models; note part of LMIC trends were cut off for visibility. LIC+ countries (ISO3): Burundi (BDI), Burkina Faso (BFA), Malawi (MWI), Nepal (NPL), Rwanda (RWA), Togo (TGO), Tajikistan (TJK), and Tanzania (TZA).

The per capita health spending trends of LIC+ and LIC-were significantly different (p < 0·0001); LIC+ and LIC-started with a notable difference in total health spending per capita (US$19 and US$34, respectively), but reached similar levels (US$44 and US$43, respectively).

The per capita government health spending trends of LIC+ and LIC-were significantly different (p < 0·0001), increasing for LIC+ and decreasing for LIC-. The average yearly rate of change in the government health spending per capita was US$0·42 for LIC+ versus US$-0·24 for LIC-.

The per capita private health spending trends of LIC+ was significantly lower than that of LIC- (p < 0·012).

### DAH on newborn & child health

Figure 4 shows the DAH per birth on newborn and childhood vaccines trends for country groups LIC- and LMIC (obtained by the mixed-effects models) and for each country in LIC+. During 2000–17, the trend of DAH per birth on vaccines of LIC+ was slightly above LIC-; the difference was not statistically significant (p > 0·19). The average annual DAH per birth was US$18·42 and US$14·01 for LIC+ and LIC-, respectively.

**Figure 4:**
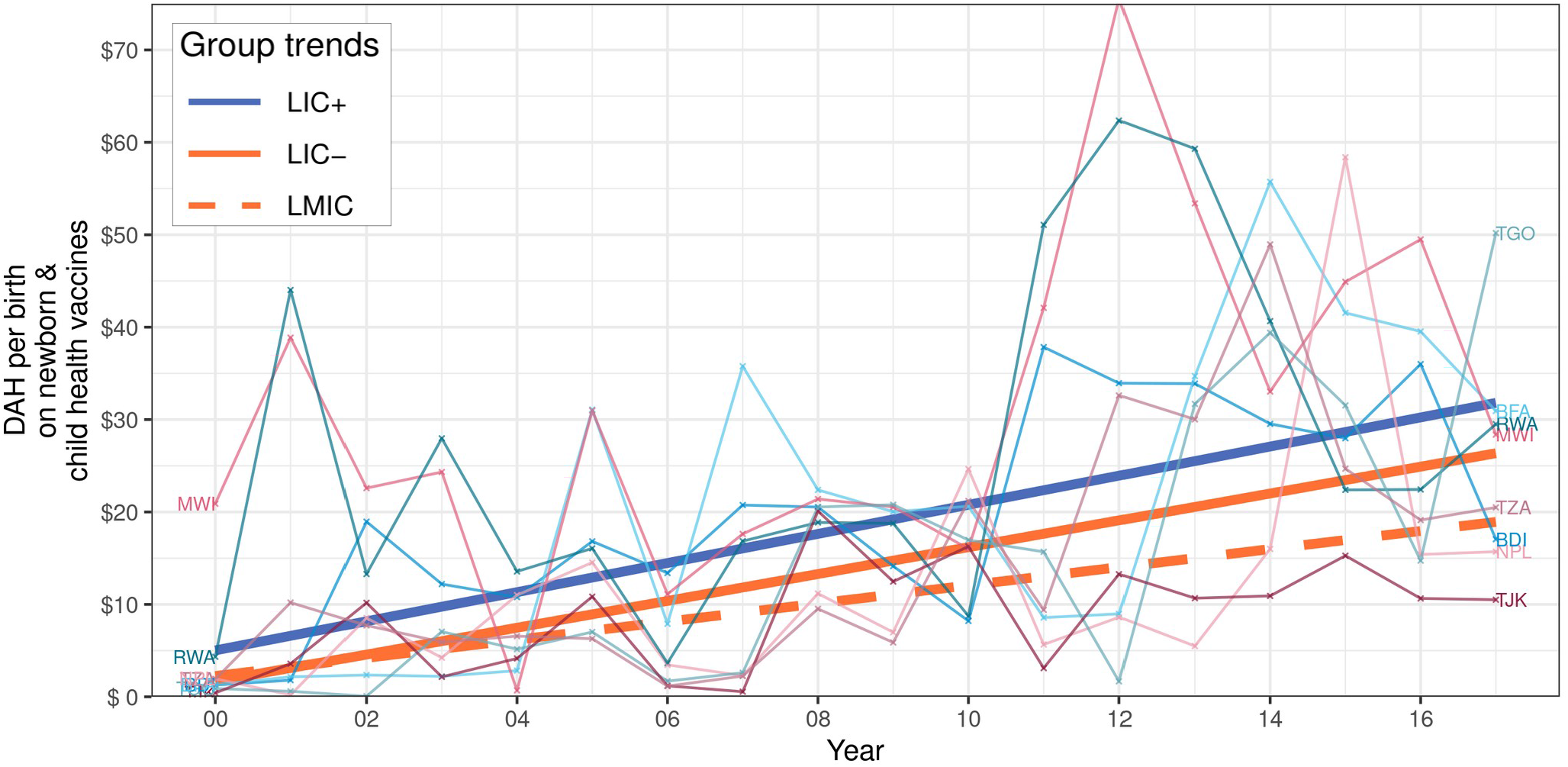
DAH per birth on newborn & child health vaccines of country groups. Data source: IHME2. The trends of LIC+, LIC-, and LMIC were fitted by linear mixed-effects models. LIC+ countries (ISO3): Burundi (BDI), Burkina Faso (BFA), Malawi (MWI), Nepal (NPL), Rwanda (RWA), Togo (TGO), Tajikistan (TJK), and Tanzania (TZA).

Figure 2 in Supplemental Materials shows the aggregated per capita DAH trends for country groups LIC- and LMIC (obtained by the mixed-effects models) and for each country in LIC+ during 2000–16. DAH per capita of LIC+ and LIC-were not significantly different (p > 0·65). LMIC had a significantly lower DAH per capita (all p < 0·0001), but a significantly higher total, government, and private health spending compared to LIC+ and LIC-.

### Routine immunization spending

Figure 5 shows the per birth total spending and government spending on routine immunization vaccines trends for country groups LIC- and LMIC (obtained by the mixed-effects models) and for each country in LIC+ from 2006 to 2017. LMIC had the highest total spending per birth on routine immunization vaccines. LIC+ spent slightly less than LMIC (difference was not significant, p > 0·73) and more than LIC- (difference was not significant, p > 0·17); there was a high variability in spending between countries and over time. For government health spending on routine immunization vaccines per birth, LIC+ had a significantly lower spending than LMIC (p < 0·0075); LIC+ and LIC-had similar trends (p > 0·53).

**Figure 5:**
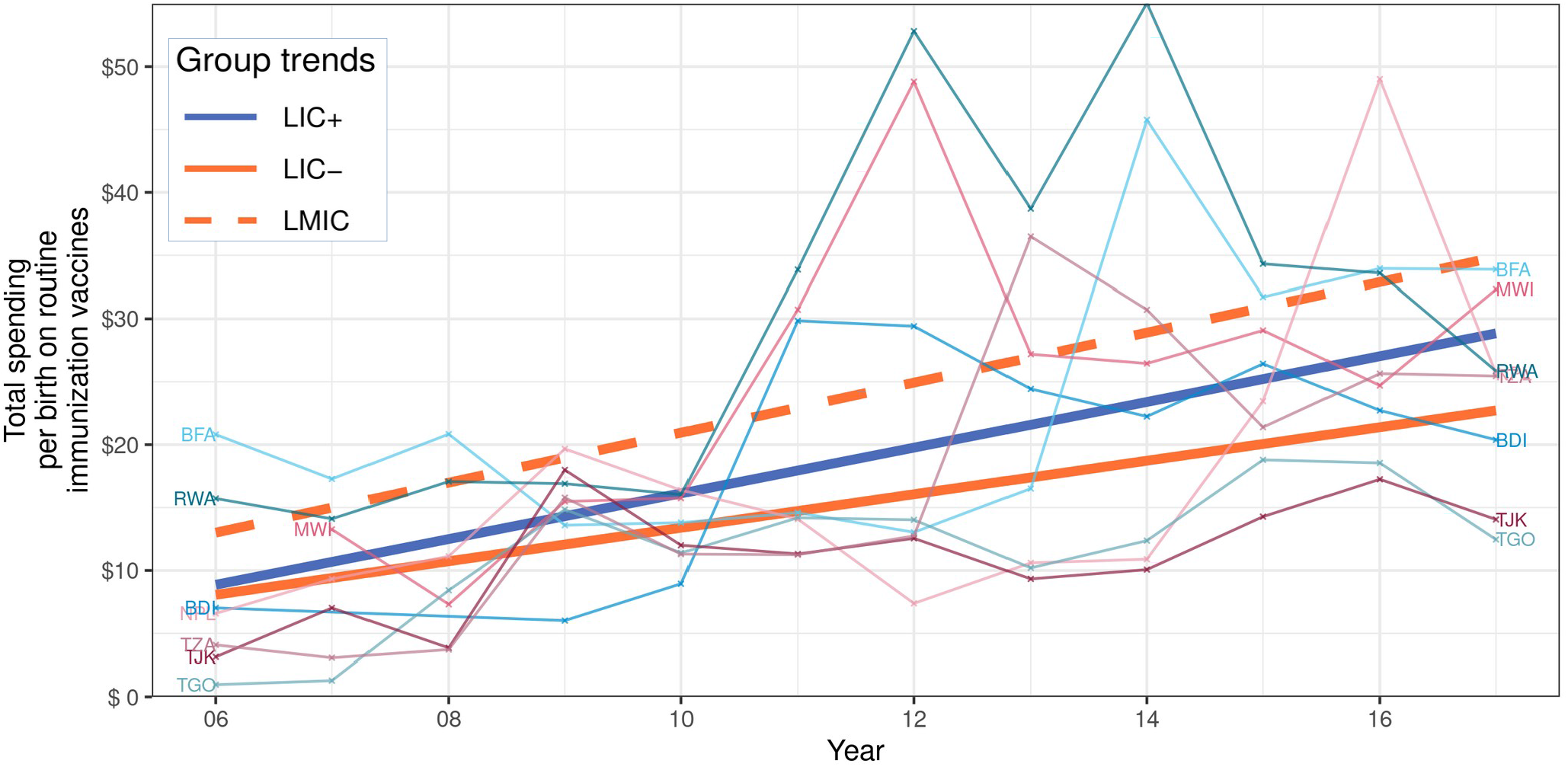
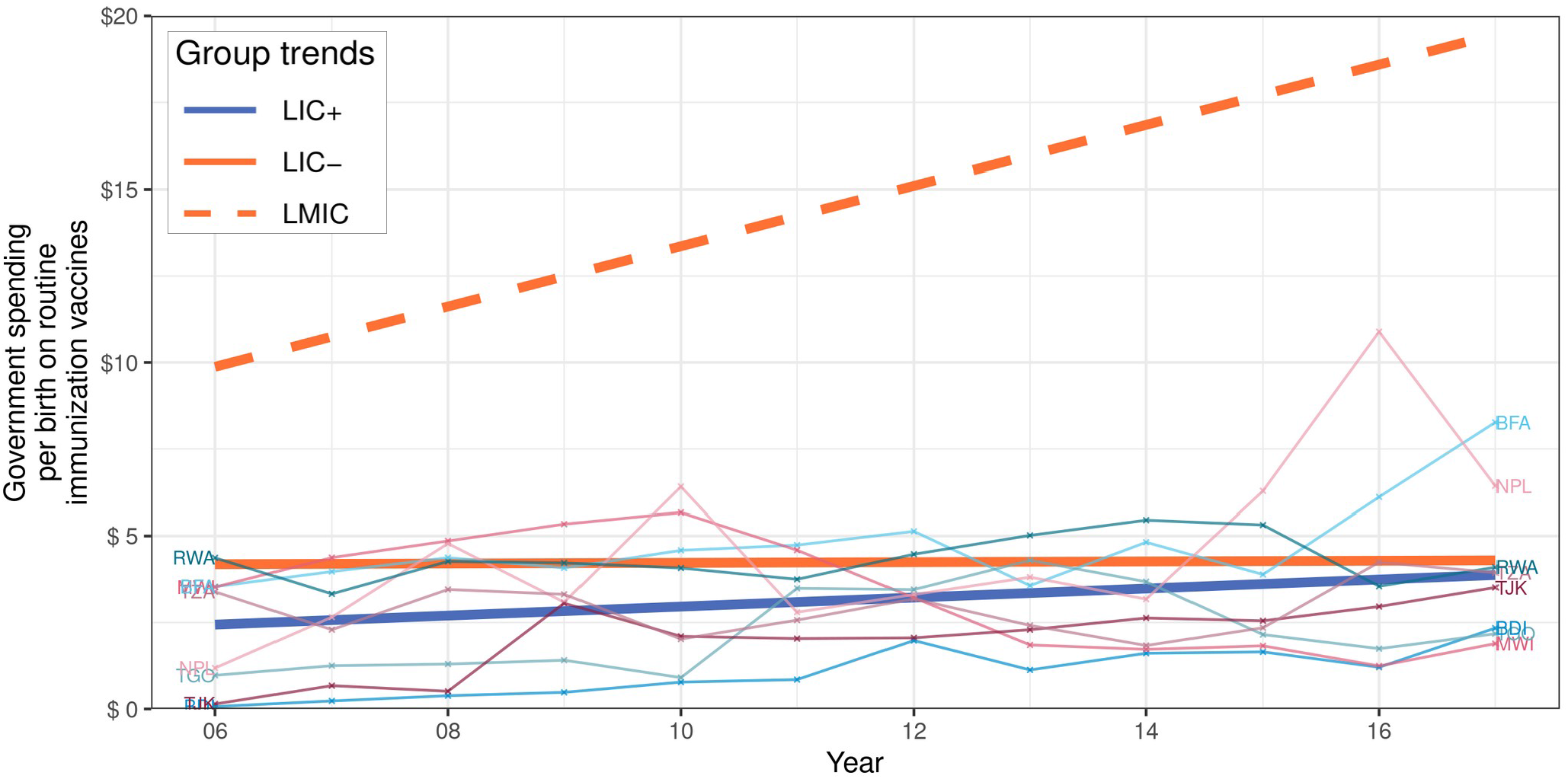
Total and government spending per birth on routine immunization vaccines of country groups. Data source: WU2. The trends of LIC+, LIC-, and LMIC were fitted by linear mixed-effects models. LIC+ countries (ISO3): Burundi (BDI), Burkina Faso (BFA), Malawi (MWI), Nepal (NPL), Rwanda (RWA), Togo (TGO), Tajikistan (TJK), and Tanzania (TZA).

## Discussion

Our analyses revealed that the exceptional vaccination coverage of LIC+ could not be explained by the countries’ economic development, total health spending, government spending on routine immunization vaccines, nor aggregated DAH. LIC+ had lower GNI, significantly lower total health spending per capita, and slightly (but not significantly) higher DAH per capita, compared to LIC-, reaching similar health spending levels only in recent years. The trends of the government health spending per birth on routine immunization vaccines of LIC+ were lower than that of LMIC, while the trends of LIC+ and LIC-remained almost constant over time and were indistinguishable from each other. Despite being considerably behind LMIC in economic development and health spending, LIC+ achieved significantly better vaccination coverage as a group. Other studies have also observed that higher spending does not always results in improved health services or outcomes^24^.

Government and private health spending per capita partly explain the vaccination performance of LIC+. The government health spending per capita of LIC+ increased over time while it decreased for LIC-; all countries in LIC+ reached or exceeded the government health spending trend of LIC-by 2016. By contrast, private health spending for LIC+ was significantly lower than that of LIC- and LMIC. These observations highlight the association between low out-of-pocket spending combined with higher government spending and higher national vaccination coverage^25^. Our study also underscores the importance of the LIC+ governments’ commitment to improving vaccination coverage, child health, and healthcare in general, making vaccines and healthcare accessible to more people. For example, previous research found that the success of Rwanda’s vaccine program was multi-factorial, where one of the main factors was a strong and high-level political will^26^. Malawi achieved the Millennium Development Goal for child survival by 2013, as their government adopted evidence-based policies and implemented programs at scale to prevent child deaths^27^. Nepal was the first LIC to have a national newborn strategy, influencing similar strategies in other countries; this was made possible due to political commitment that supported newborn survival^28^.

DAH on vaccines and child health also partly explain the difference in vaccination performance between LIC+ and LIC-; although the evidence is less conclusive than for the previously mentioned indicators. LIC+ received slightly more DAH per birth on newborn and child health vaccines than LIC-, but these trends were not statistically different, possibly in part due to the following two factors: (i) the large variation in DAH on vaccines year-to-year or across the LIC+ countries, and (ii) DAH funding that was used for purposes different to routine immunization such as introduction of new vaccines, health system strengthening, and supplementary activities. For example, the introductions of the pneumococcal vaccine in Malawi in November 2011^29^ and Nepal in January 2015^30^, were associated with the increase of DAH on newborn and child health during those years. Previous studies have suggested that LIC will remain dependent on DAH in the near future, unless they increase government health spending significantly^11^.

In summary, our analysis suggests that government health spending, with a high-level political will and newborn focused programs, and DAH on vaccines and child health may have led to lower private health spending, efficient utilization of healthcare resources, and the immunization success of LIC+. The financial commitment of LIC+ governments was clear as their health spending increased over time, as opposed to LIC-that decreased/stagnated. Agencies that actively invest into countries or programs to improve vaccination coverage might want to consider the countries’ government and private health spending levels when making investment decisions. LIC continue to be dependent on DAH to achieve high vaccination coverage and remain far behind in government health spending on vaccines when compared to LMIC.

## Data Availability

All data used in the study is publicly available as described in the paper.

https://data.unicef.org/resources/dataset/immunization/

https://data.worldbank.org

http://ghdx.healthdata.org/record/ihme-data/global-health-spending-1995-2016

http://ghdx.healthdata.org/record/ihme-data/development-assistance-health-database-1990-2018

https://www.who.int/immunization/programmes_systems/financing/data_indicators/en/

## Acknowledgments

This work was supported by The Bill & Melinda Gates Foundation (OPP1191549). This research has also been supported in part by the William W. George endowment and the following benefactors at Georgia Tech: Andrea Laliberte, Joseph C. Mello, Richard E. & Charlene Zalesky, and Claudia & Paul Raines. We are grateful to Kyra Hester, Robert Bednarczyk, David Phillips, Gloria Ikilezi, and Anna Rapp for feedback and discussion that helped with the content and exploration of this document.

