## Supplemental materials for "Health spending and vaccination coverage in low-income countries"

*Francisco Castillo-Zunino[a] – Pinar Keskinocak[a] – Dima Nazzal[a] – Matthew C Freeman[b].*
*[a] H. Milton Stewart School of Industrial and Systems Engineering, Georgia Institute of Technology, Atlanta, GA, USA.*
*[b] Rollins School of Public Health, Emory University, Atlanta, GA, USA.*

| Data source | | Variable | Missing data % | Time period |
|---|---|---|---|---|
| Short | Full | | | |
| WU1[1] | WHO-UNICEF: estimates of national infant immunization coverage | DTP1 coverage | 0 | 2000–18 |
| | | DTP3 coverage | 0 | |
| | | MCV1 coverage | 0 | |
| | | BCG coverage | 0 | |
| | | Pol3 coverage | 0 | |
| WB1[2] | World Bank: world development indicators | Population | 0 | 2000–17 |
| | | Birth rate | 0 | |
| | | GNI per capita | 2·1 | 2000–18 |
| | | GDP per capita | 1·3 | |
| IHME1[3] | IHME: global health spending 1995–2016 | Total health spending per capita | 0 | 2000–16 |
| | | Government health spending per capita | 0 | |
| | | Out-of-pocket health spending per capita | 0 | |
| | | Prepaid private health spending per capita | 0 | |
| | | DAH per capita | 0 | |
| IHME2[4*] | IHME: development assistance for health database 1990–2018 | DAH per birth on newborn & child health | 0 | 2000–17 |
| | | DAH per birth on newborn & child health vaccines | 0 | |
| WU2[5*] | WHO-UNICEF joint reporting form: immunization financing indicators | Total spending per birth on routine immunization | 19·6 | 2006–17 |
| | | Government spending per birth on routine immunization | 15·1 | |
| | | Total spending per birth on routine immunization vaccines | 6·6 | |
| | | Government spending per birth on routine immunization vaccines | 5·4 | |

*Table 1: Summary of data sources*

** With data source WB1 we calculated per birth values (using population and live birth rate).*

| **Indicator (US$)** | **Year range** | **Starting trend value** | | | **Ending trend value** | | | **Yearly change rate** | | |
|---|---|---|---|---|---|---|---|---|---|---|
| | | **LIC+** | **LIC-** | **LMIC** | **LIC+** | **LIC-** | **LMIC** | **LIC+** | **LIC-** | **LMIC** |
| GNI per capita | 2000–18 | 199 | 320 | 571 | 785 | 831 | 2487 | 32·59 | 28·43 | 106·42 |
| GDP per capita | | 217 | 351 | 658 | 765 | 859 | 2576 | 30·43 | 28·25 | 106·57 |
| Total health spending per capita | 2000–16 | 18·64 | 34·47 | 62·75 | 43·87 | 43·41 | 110·55 | 1·58 | 0·56 | 2·99 |
| Government health spending per capita | | 4·30 | 10·83 | 23·51 | 11·05 | 7·02 | 47·98 | 0·42 | -0·24 | 1·53 |
| Private health spending per capita✢ | | 10·67 | 21·06 | 35·55 | 17·43 | 23·29 | 53·96 | 0·42 | 0·14 | 1·15 |
| DAH per capita | | 3·87 | 2·58 | 3·71 | 15·35 | 13·07 | 8·42 | 0·72 | 0·66 | 0·29 |
| DAH per birth on newborn & child health | 2000–17 | 1·23 | 2·74 | 11·61 | 81·62 | 67·81 | 51·85 | 4·73 | 3·83 | 2·37 |
| DAH per birth on newborn & child health vaccines | | 5·03 | 1·67 | 2·25 | 31·81 | 26·36 | 18·93 | 1·58 | 1·45 | 0·98 |
| Total spending per birth on routine immunization | 2006–17 | 15·68 | 11·55 | 25·25 | 37·15 | 31·54 | 45·15 | 1·95 | 1·82 | 1·81 |
| Government spending per birth on routine immunization | | 4·34 | 4·59 | 16·17 | 7·95 | 6·74 | 23·43 | 0·33 | 0·20 | 0·66 |
| Total spending per birth on routine immunization vaccines | | 8·86 | 8·06 | 13·00 | 28·85 | 22·72 | 34·87 | 1·82 | 1·33 | 1·99 |
| Government spending per birth on routine immunization vaccines | | 2·44 | 4·19 | 9·87 | 3·87 | 4·30 | 19·46 | 0·13 | 0·01 | 0·87 |

*Table 2: Summary of financial trends of country groups*

*Each indicator was fitted by a linear mixed-effects model. The table shows the values of the trends in the first and last years (intercepts at different times), and the yearly change rate over time (slope).*
*✢ Private health spending is the sum of out-of-pocket and prepaid private health spending.*

| Indicator (US$) | LIC+ & LIC- comparison | | | LIC+ & LMIC comparison | | |
| --- | --- | --- | --- | --- | --- | --- |
| | **$\chi^2$** | **KR** | **PB** | **$\chi^2$** | **KR** | **PB** |
| GNI per capita | 0·1325 | 0·1502 | 0·1468 | <0·0001* | <0·0001* | <0·0001* |
| GDP per capita | 0·4155 | 0·4404 | 0·4403 | <0·0001* | <0·0001* | <0·0001* |
| Total health spending per capita | <0·0001* | <0·0001* | <0·0001* | <0·0001* | <0·0001* | <0·0001* |
| Government health spending per capita | <0·0001* | <0·0001* | <0·0001* | <0·0001* | <0·0001* | <0·0001* |
| Private health spending per capita✢ | 0·0071* | 0·0116* | 0·0097* | <0·0001* | <0·0001* | <0·0001* |
| DAH per capita | 0·6537 | 0·6734 | 0·6663 | <0·0001* | <0·0001* | <0·0001* |
| DAH per birth on newborn & child health | 0·0794 | 0·0937 | 0·0898 | <0·0001* | <0·0001* | <0·0001* |
| DAH per birth on newborn & child health vaccines | 0·1892 | 0·2104 | 0·2058 | <0·0001* | 0·0002* | 0·0002* |
| Total spending per birth on routine immunization | 0·3981 | 0·4293 | 0·4199 | 0·6131 | 0·6284 | 0·6257 |
| Government spending per birth on routine immunization | 0·5827 | 0·5929 | 0·6033 | 0·1360 | 0·1474 | 0·1502 |
| Total spending per birth on routine immunization vaccines | 0·1704 | 0·1920 | 0·1898 | 0·7276 | 0·7381 | 0·7365 |
| Government spending per birth on routine immunization vaccines | 0·5286 | 0·5419 | 0·5479 | 0·0055* | 0·0075* | 0·0072* |

*Table 3: Significance testing financial trends of LIC+ compared to LIC- and LMIC*

*χ2, KR, and PB represent the p-values of an asymptotic $\chi^2$ test, a Kenward-Roger approximation for F tests for reduction of mean structure, and a parametric bootstrap method (10 000 simulations) respectively[6]. When a p-value is significant it means the group trends are significantly different to each other; i.e., it does not refer to the significance of a specific parameter of the regression but to the significance of all parameters combined.*

✢ *Private health spending is the sum of out-of-pocket and prepaid private health spending.*

* *p-values are significant with $p < 0·05$.*

### Additional plots

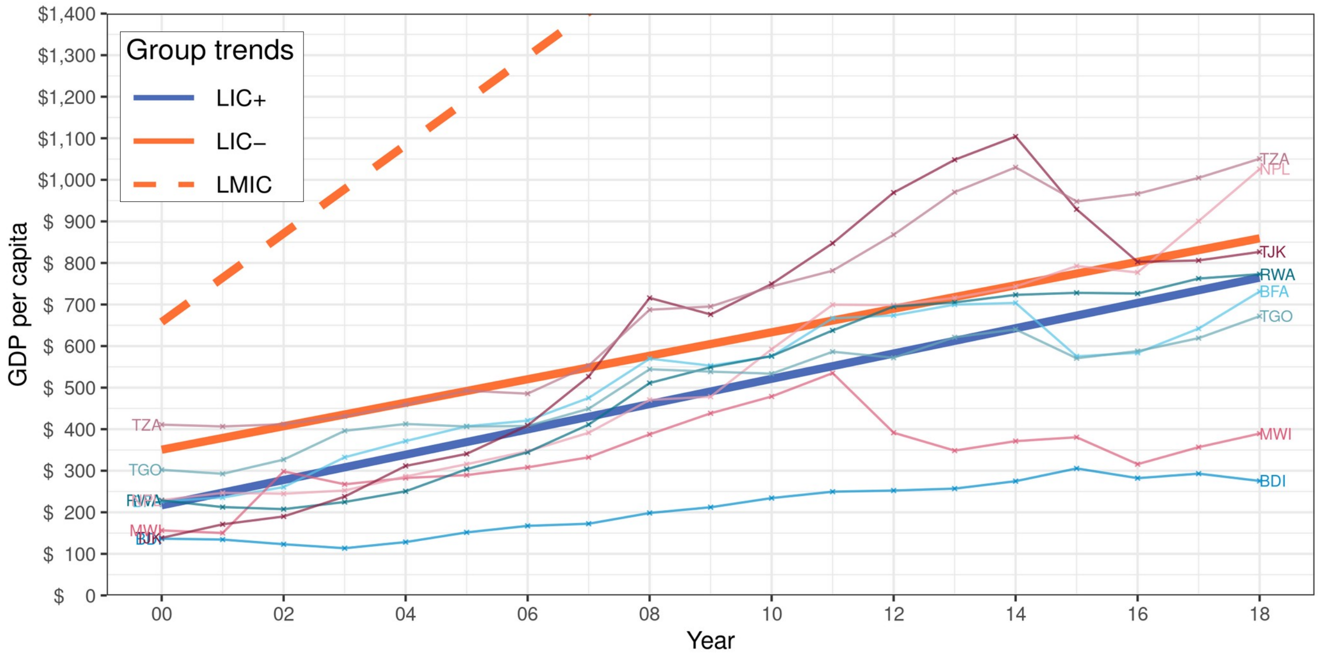


*Figure 1: GDP per capita of country groups*

*Data source: WB1. The trends of LIC+, LIC-, and LMIC were fitted by linear mixed-effects models; note part of the LMIC trend was cut off for visibility. LIC+ countries (ISO3): Burundi (BDI), Burkina Faso (BFA), Malawi (MWI), Nepal (NPL), Rwanda (RWA), Togo (TGO), Tajikistan (TJK), and Tanzania (TZA).*

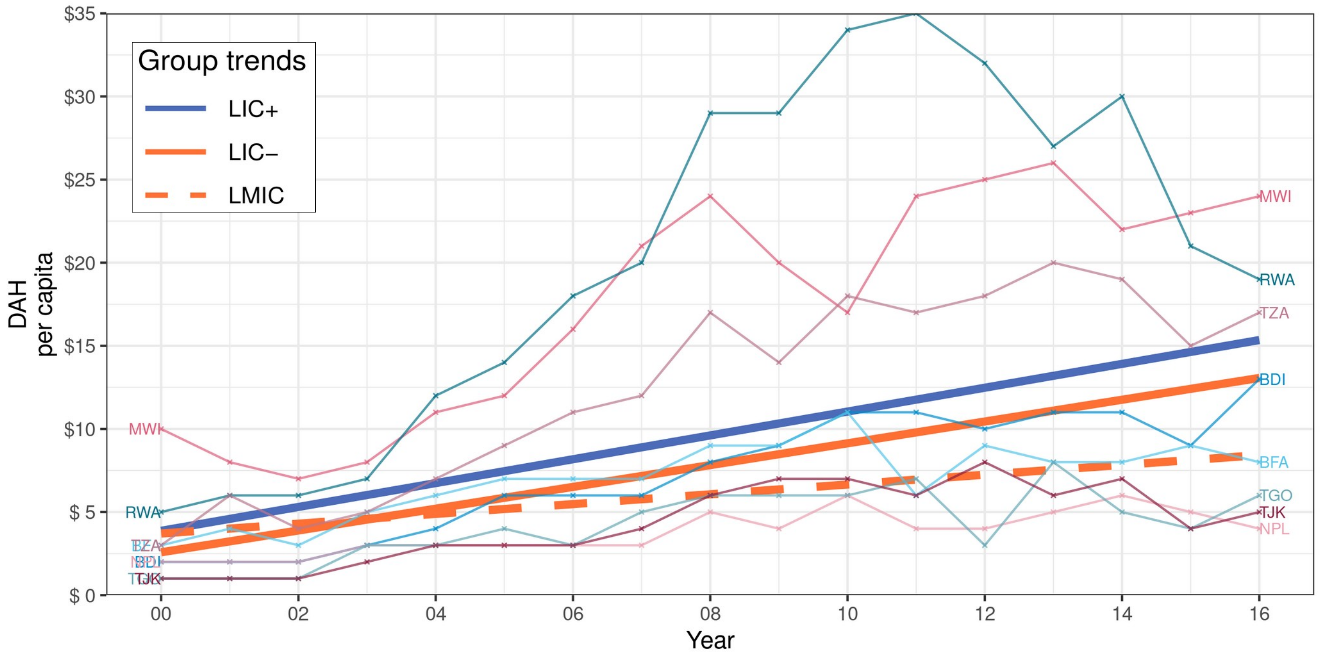


Figure 2: DAH per capita of country groups

*Data source: IHME1. The trends of LIC+, LIC-, and LMIC were fitted by linear mixed-effects models. LIC+ countries (ISO3): Burundi (BDI), Burkina Faso (BFA), Malawi (MWI), Nepal (NPL), Rwanda (RWA), Togo (TGO), Tajikistan (TJK), and Tanzania (TZA).*

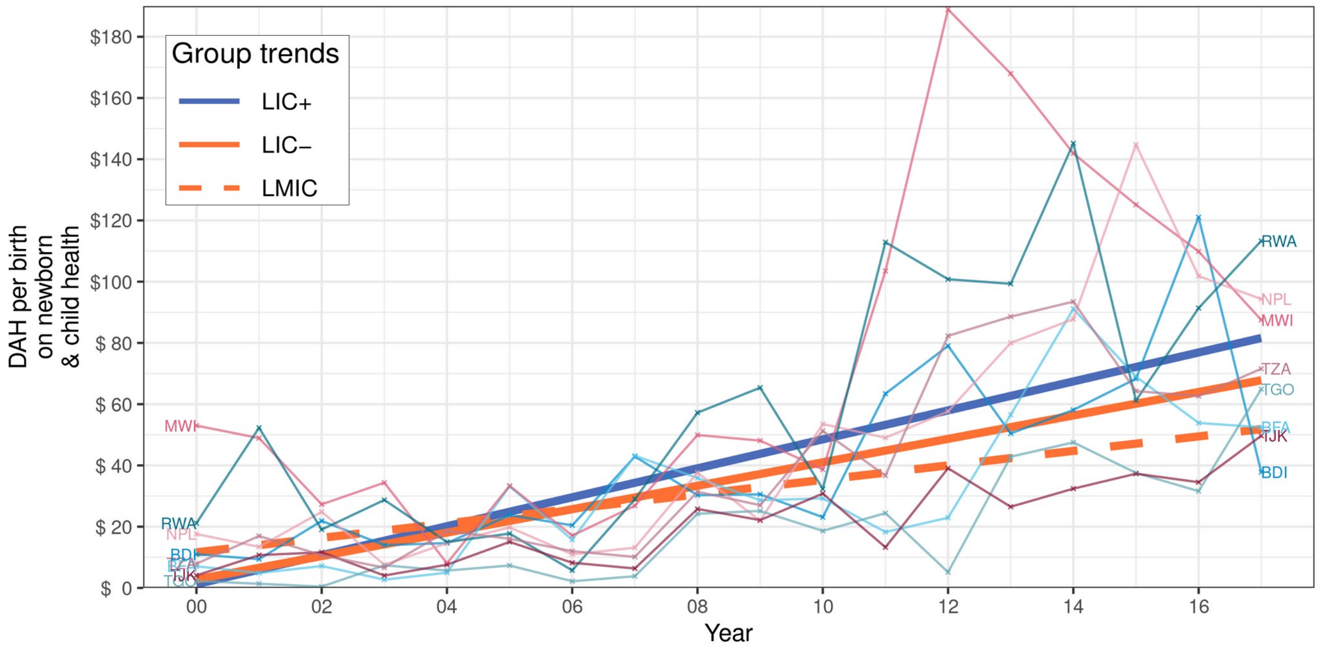


Figure 3: DAH per birth on newborn & child health of country groups

*Data source: IHME1. The trends of LIC+, LIC-, and LMIC were fitted by linear mixed-effects models. LIC+ countries (ISO3): Burundi (BDI), Burkina Faso (BFA), Malawi (MWI), Nepal (NPL), Rwanda (RWA), Togo (TGO), Tajikistan (TJK), and Tanzania (TZA).*

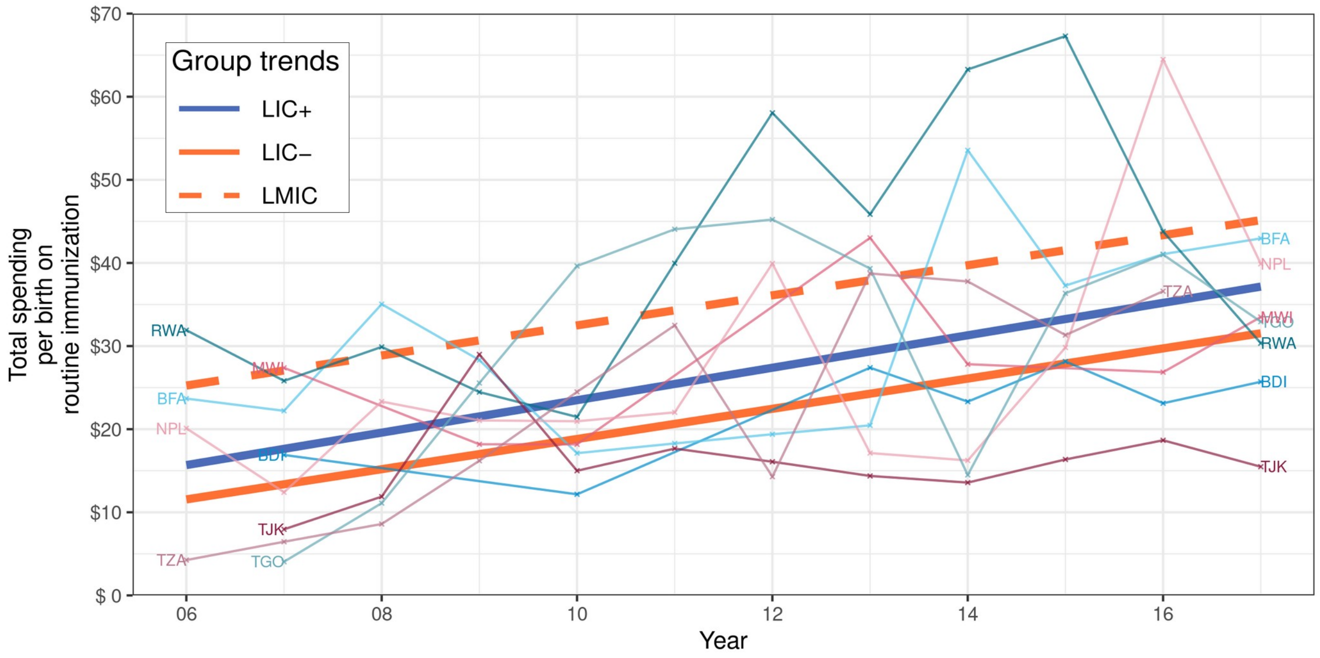


Figure 4: Total spending per birth on routine immunization of country groups

*Data source: IHME1. The trends of LIC+, LIC-, and LMIC were fitted by linear mixed-effects models. LIC+ countries (ISO3): Burundi (BDI), Burkina Faso (BFA), Malawi (MWI), Nepal (NPL), Rwanda (RWA), Togo (TGO), Tajikistan (TJK), and Tanzania (TZA).*

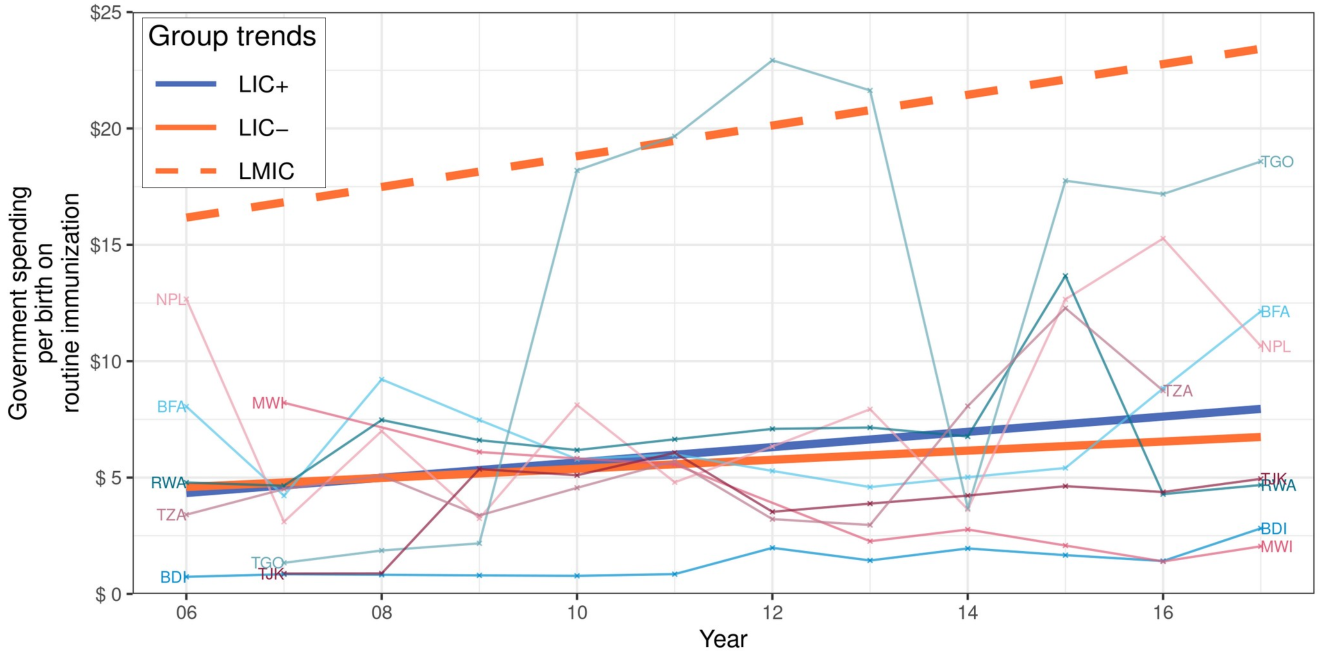


Figure 5: Government spending per birth on routine immunization of country groups
Data source: IHME1. The trends of LIC+, LIC-, and LMIC were fitted by linear mixed-effects models.
*LIC+ countries (ISO3)*: Burundi (BDI), Burkina Faso (BFA), Malawi (MWI), Nepal (NPL), Rwanda (RWA), Togo (TGO), Tajikistan (TJK), and Tanzania (TZA).

### Mixed-effects models

#### Model definition

We implemented all linear mixed-effects models with the R library *lme4*[7] by running the code

$$lmer(var \sim year * group + (1|country), data = sample) \quad (1)$$

where *var* represents the variable being fitted over time, *sample* represents the set with all data points, and the rest represent the *year*, *country*, and *group* (LIC+, LIC- or LMIC) of the data points. The models used can be formulated as follows:

$$var_{tj} = \beta_{0j} + t\beta_{1j} + R_{tj} \quad \text{for each year } t \text{ and country } j \quad (2)$$

$$\beta_{0j} = \alpha_{00} + U_j \quad \text{for each country } j \text{ in LIC+} \quad (3)$$

$$\beta_{0j} = \alpha_{00} + \alpha_{01} + U_j \quad \text{for each country } j \text{ in LIC-} \quad (4)$$

$$\beta_{0j} = \alpha_{00} + \alpha_{02} + U_j \quad \text{for each country } j \text{ in LMIC} \quad (5)$$

$$\beta_{1j} = \alpha_{10} \quad \text{for each country } j \text{ in LIC+} \quad (6)$$

$$\beta_{1j} = \alpha_{10} + \alpha_{11} \quad \text{for each country } j \text{ in LIC-} \quad (7)$$

$$\beta_{1j} = \alpha_{10} + \alpha_{12} \quad \text{for each country } j \text{ in LMIC} \quad (8)$$

$$R_{tj} \sim N(0, \sigma^2) \quad \text{for each year } t \text{ and country } j \quad (9)$$

$$U_j \sim N(0, \tau^2) \quad \text{for each country } j \quad (10)$$

Equation (2) represents the linear regression of the tested variable dependent of time. The data value in year $t$ of country $j$ is represented by $var_{tj}$. Coefficients $\beta_{0j}$ and $\beta_{1j}$ are the intercept and slope respectively of each country $j$, which changes depending on which group they belong to. Random variables $R_{tj}$ illustrate the random noise within samples, that have a normal distribution with mean 0 and variance $\sigma^2$ as shown in equation (9).

Equations (3) to (5) represent the intercept coefficients of LIC+, LIC-, and LMIC respectively. LIC+ have a base intercept $\alpha_{00}$, then each LIC- and LMIC have their own differences from the base intercept (coefficients $\alpha_{01}$ and $\alpha_{02}$ respectively). The starting year t = 0 can be set to any year from 2000 to 2018, depending where we wanted to test the intercept values of groups.

Equations (6) to (8) represent the slope coefficients of LIC+, LIC-, and LMIC respectively. LIC+ have a base slope $\alpha_{10}$ , then each LIC- and LMIC have their own differences from the base slope (coefficients $\alpha_{11}$ and $\alpha_{12}$ respectively). Random variables $U_j$ consider intercept variations within each country; they have a normal distribution with mean 0 and variance $\tau^2$ as seen in equation (10).

### Significance testing

We performed significance testing between specific country groups: LIC+ compared to LIC-, and LIC+ compared to LMIC. We used the following code in R, with library *pbkrtest*[6], to test the differences:

$$model = lmer(var \sim year * group.binary + (1|country), data = sample.binary\ ) \tag{11}$$

$$model.no.group = lmer(var \sim year + (1|country), data = sample.binary\ ) \tag{12}$$

$$anova(model, model.no.group) \tag{13}$$

$$KRmodcomp(model, model.no.group) \tag{14}$$

$$P\ Bmodcomp(model, model.no.group, nsim = 10000) \tag{15}$$

Line (11) shows a model almost identical to (1), the differences are that the former has group.binary which is a binary variable (either LIC+ or not), and the sample.binary only considers countries in LIC+ and the other compared group (not all three groups together as in (1)). Line (12) shows a reduced version of the model in (11), with no grouping binary variable. Both models in (11) and (12) are compared in lines (13), (14), and (15) by computing the asymptotic $\chi^2$ test, the Kenward-Roger approximation, and parametric bootstrap[6], respectively. Line (15) runs 10 000 parametric bootstrap replications to obtain a good estimate.

### Alternative grouping criteria

Consider an alternative grouping criterion to split LIC into LIC+ and LIC- to align strictly with the Global Vaccine Action Plan goal of 90% DTP3 coverage[8] as opposed to the 85% in the original criteria, and increase the dropout threshold from 5% to 10%. These adjustments to the criteria result in minor changes to the LIC+ and LIC- country composition: (i) Malawi and Togo are moved from LIC+ to the LIC- group, for having 88·4% and 88·6% DTP3 coverage respectively, and (ii) Uganda is moved from LIC- to LIC+ for having a 6·6% dropout rate. The main findings in this paper are consistent with the findings under this alternative grouping; results and plots can be seen below.

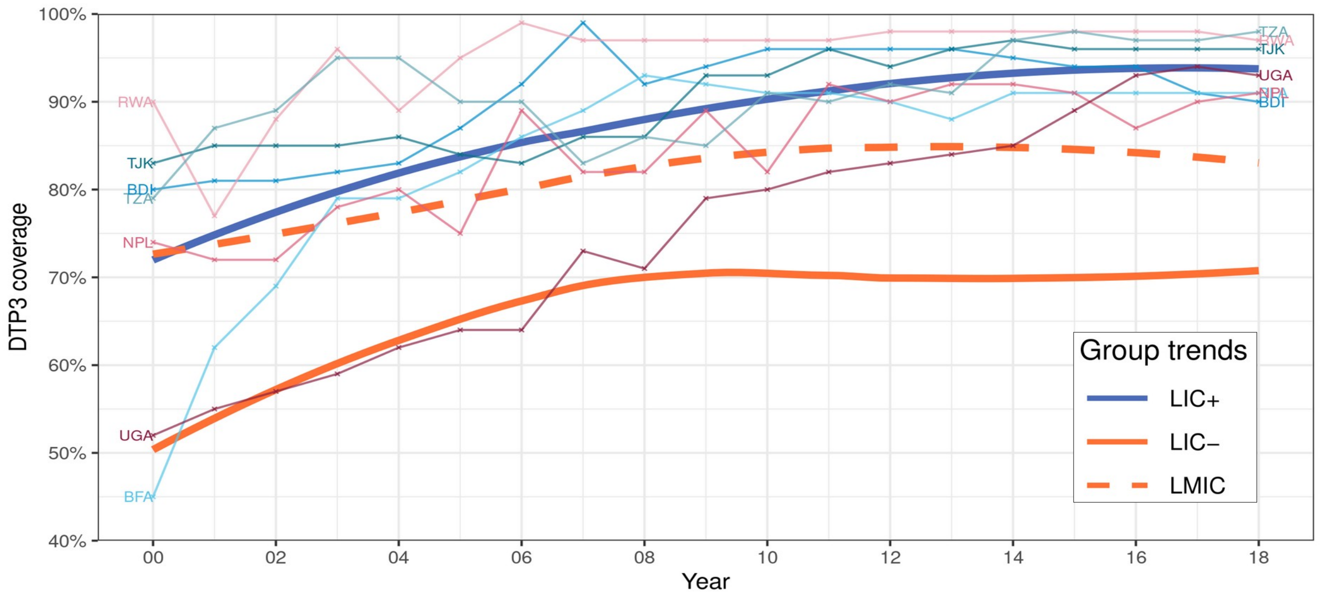


Figure 6: *DTP3* coverage of country groups – alternative country groups

*Data source: WU1. The trends of LIC+, LIC-, and LMIC were fitted by a local polynomial regression (a locally estimated scatter-plot smoothing; usually referred to as loess*[9]*). LIC+ countries (ISO3): Burundi (BDI), Burkina Faso (BFA), Nepal (NPL), Rwanda (RWA), Tajikistan (TJK), Tanzania (TZA), and Uganda (UGA).*

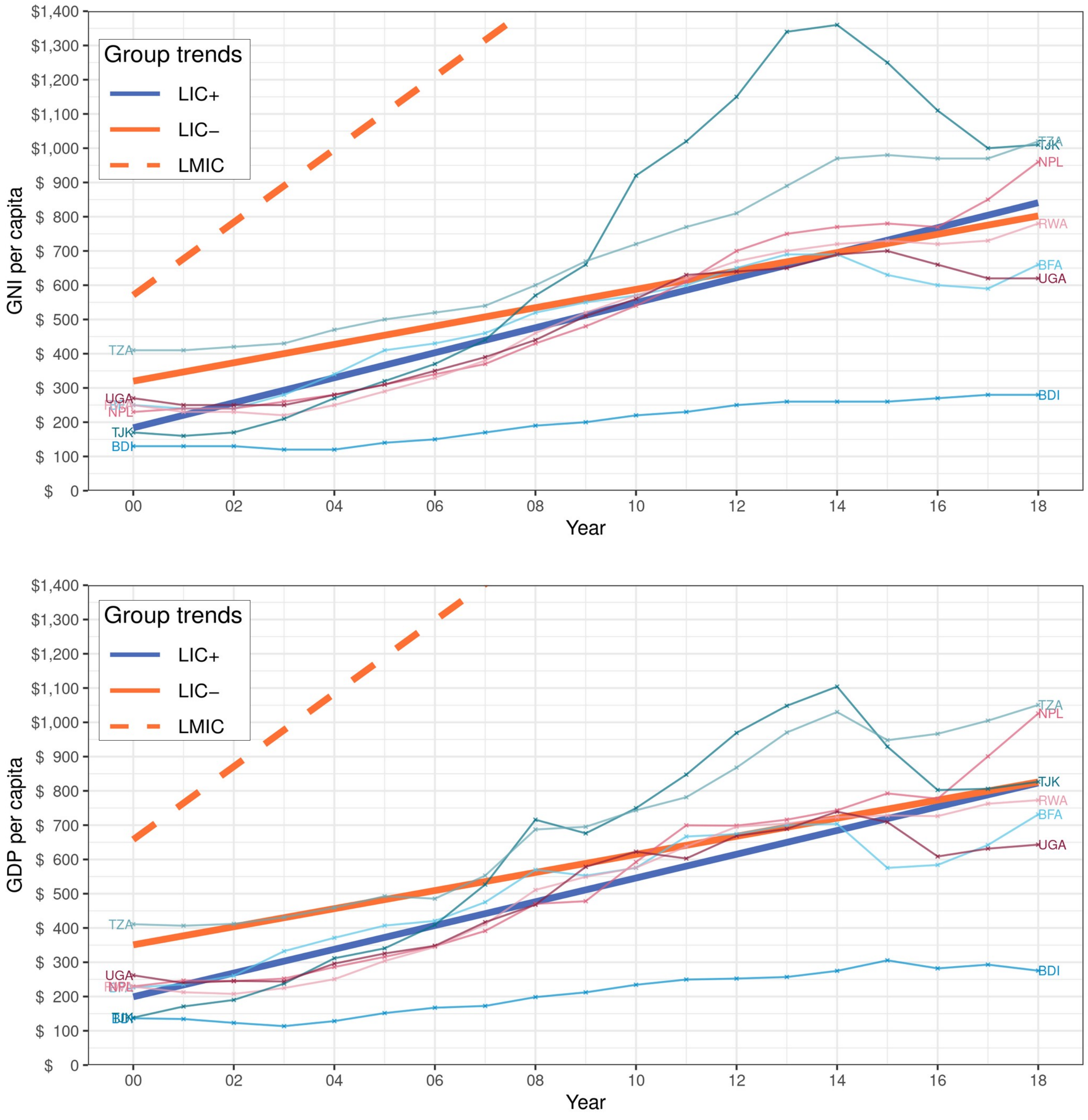


*Figure 7: GNI & GDP per capita of country groups – alternative country groups*

*Data source: WB1. The trends of LIC+, LIC-, and LMIC were fitted by linear mixed-effects models; note part of the LMIC trend was cut off for visibility. LIC+ countries (ISO3): Burundi (BDI), Burkina Faso (BFA), Nepal (NPL), Rwanda (RWA), Tajikistan (TJK), Tanzania (TZA), and Uganda (UGA).*

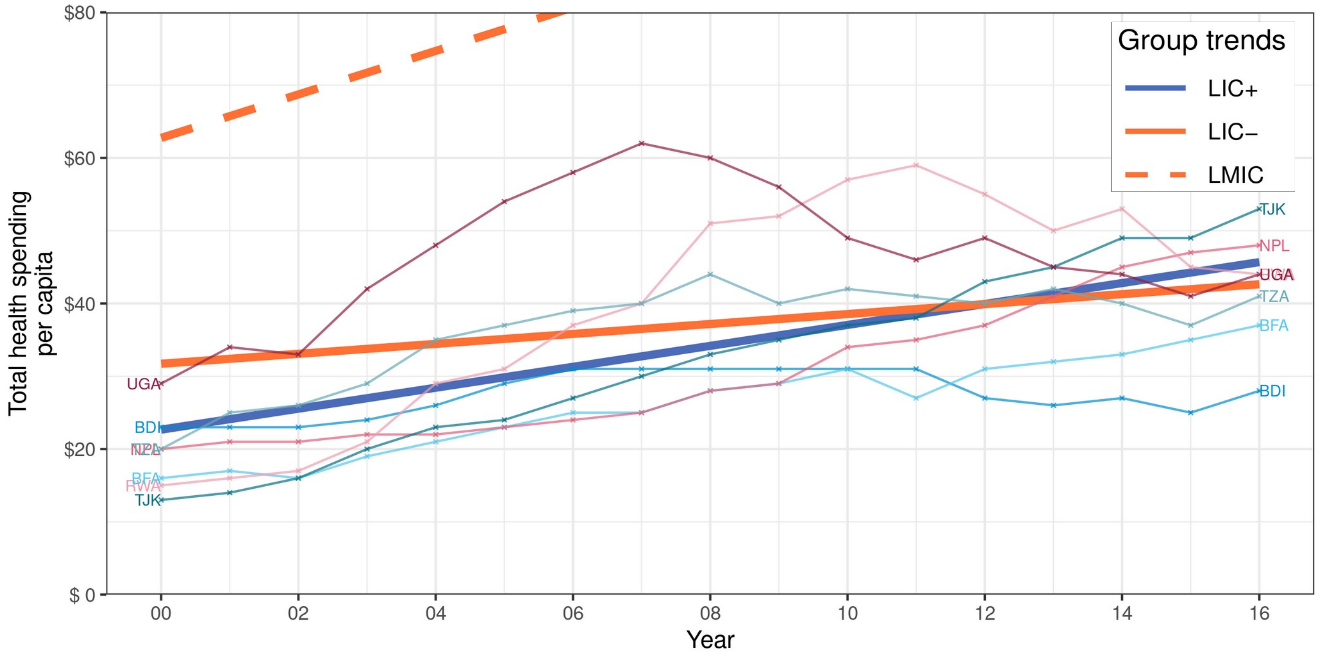
Group trends
LIC+
LIC−
LMIC
Total health spending per capita
Year
$80
$60
$40
$20
$ 0
00
02
04
06
08
10
12
14
16
UGA
BDI
NPL
BFA
RWA
TJK
TZA

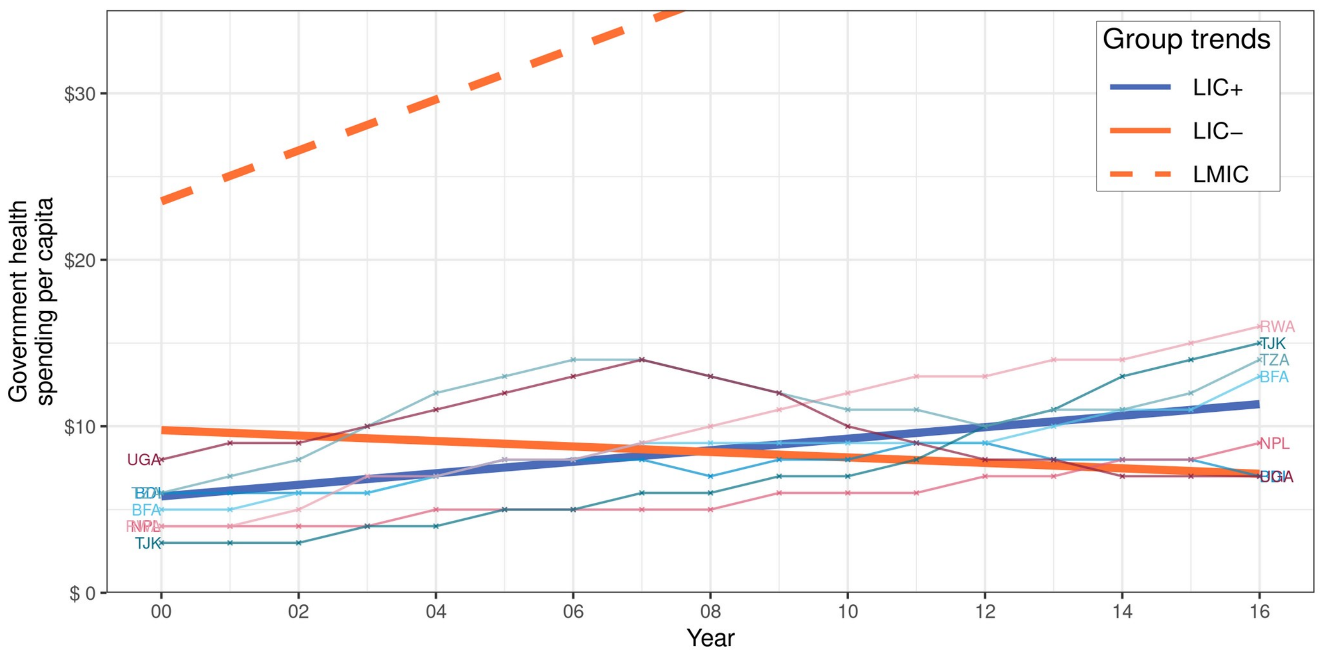
Group trends
LIC+
LIC−
LMIC
Government health spending per capita
Year
$30
$20
$10
$ 0
00
02
04
06
08
10
12
14
16
UGA
BDI
BFA
NPL
TJK
RWA
TZA

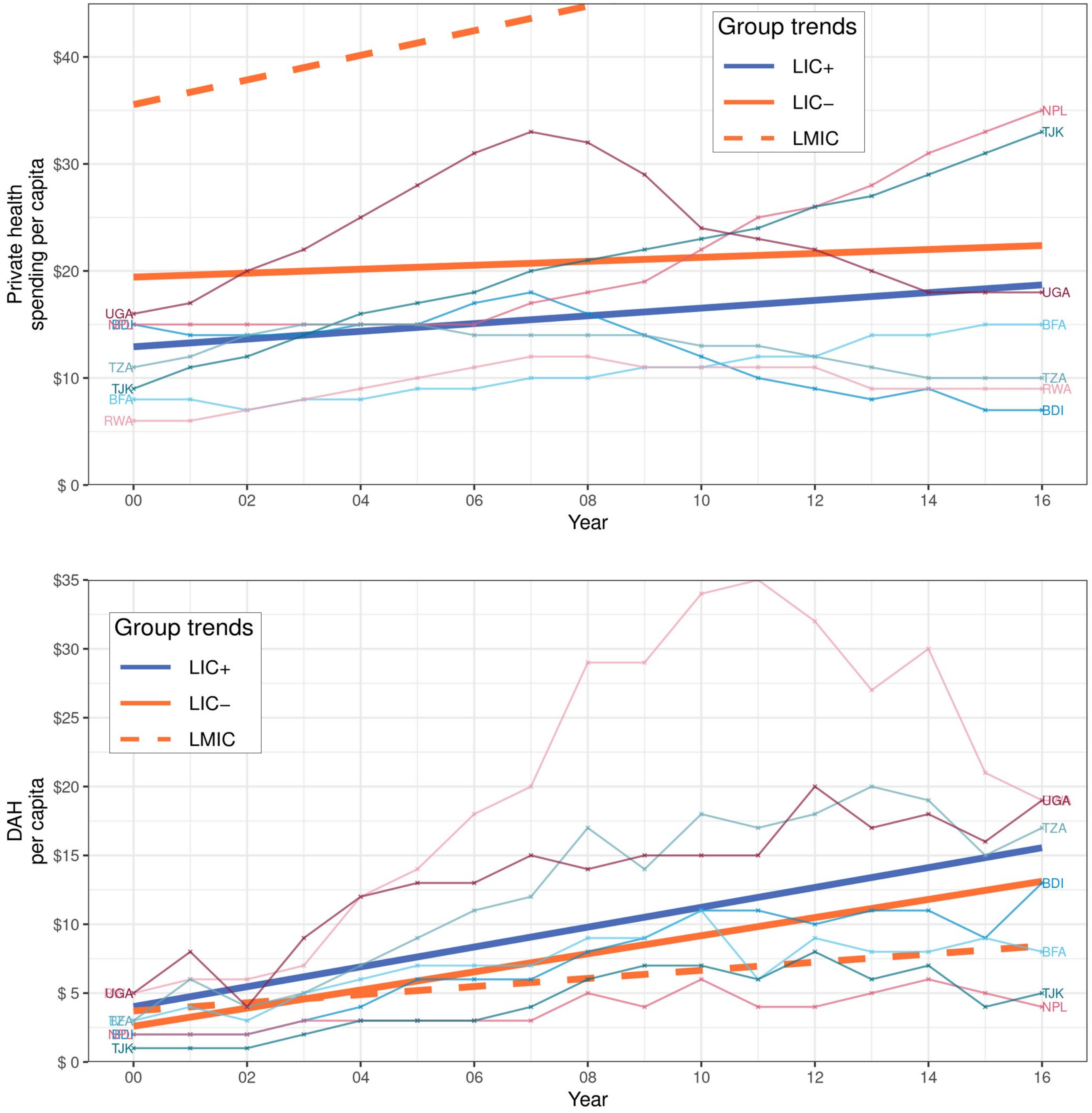


*Figure 8: Total, government, private health spending, and DAH per capita of country groups – alternative country groups*

*Data source: IHME1. The trends of LIC+, LIC-, and LMIC were fitted by linear mixed-effects models; note part of LMIC trends were cut off for visibility. LIC+ countries (ISO3): Burundi (BDI), Burkina Faso (BFA), Nepal (NPL), Rwanda (RWA), Tajikistan (TJK), Tanzania (TZA), and Uganda (UGA).*

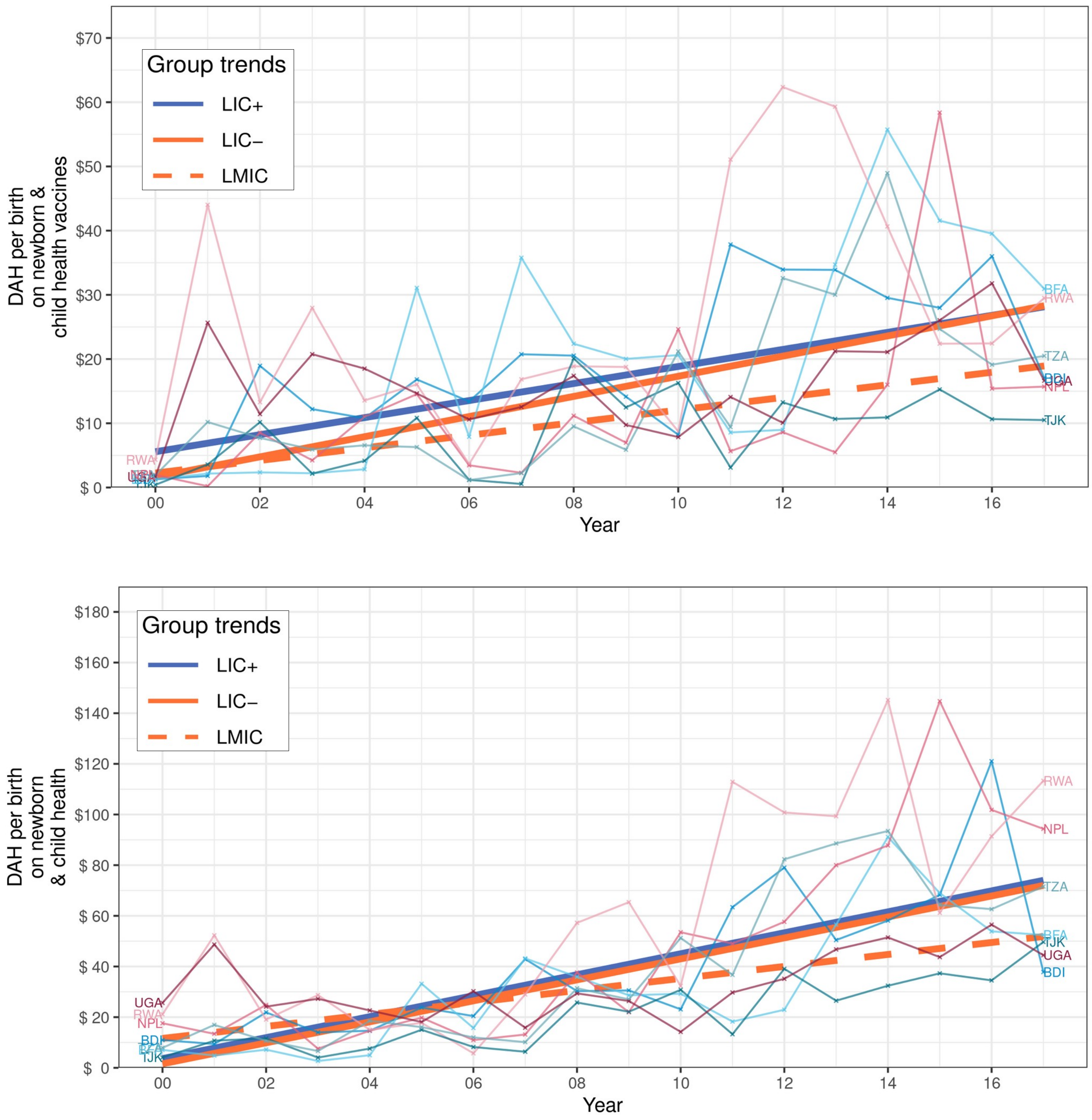


*Figure 9: DAH per birth on vaccines and on newborn & child health in general of country groups – alternative country groups*

*Data source: IHME2. The trends of LIC+, LIC-, and LMIC were fitted by linear mixed-effects models. LIC+ countries (ISO3): Burundi (BDI), Burkina Faso (BFA), Nepal (NPL), Rwanda (RWA), Tajikistan (TJK), Tanzania (TZA), and Uganda (UGA).*

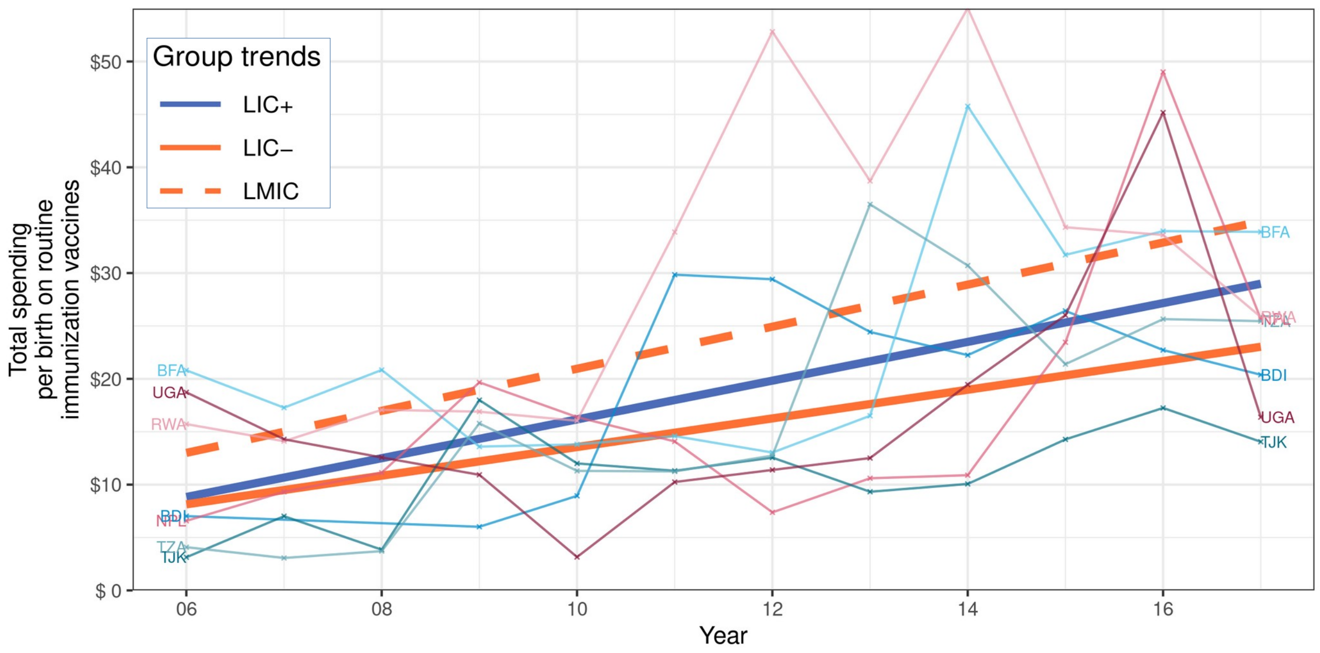


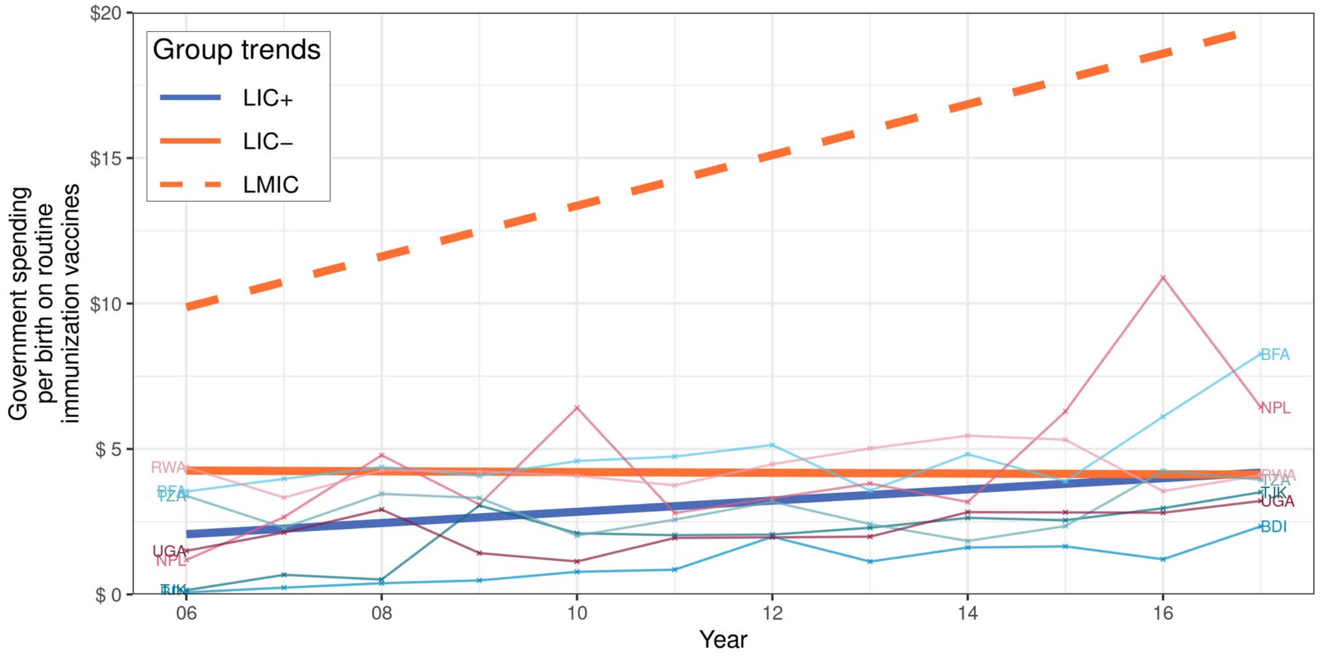


*Figure 10: Total and government spending per birth on routine immunization vaccines of country groups – alternative country group*

*Data source: WU2. The trends of LIC+, LIC-, and LMIC were fitted by linear mixed-effects models. LIC+ countries (ISO3): Burundi (BDI), Burkina Faso (BFA), Nepal (NPL), Rwanda (RWA), Tajikistan (TJK), Tanzania (TZA), and Uganda (UGA).*

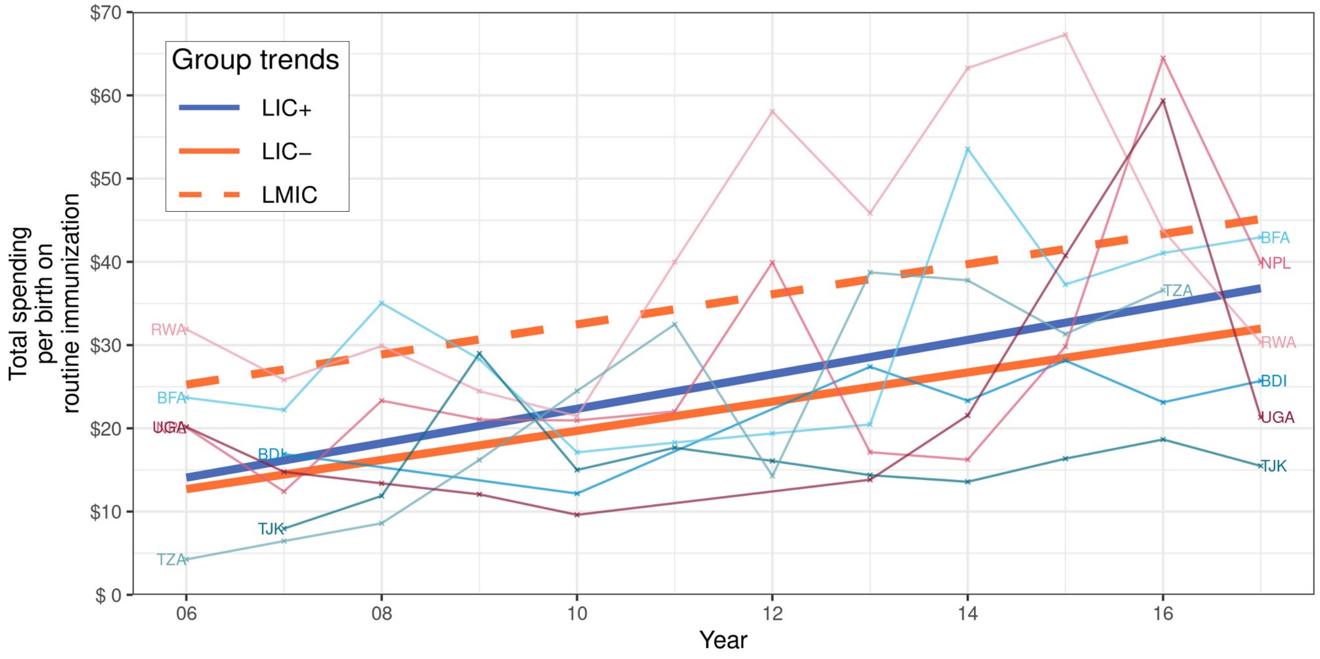


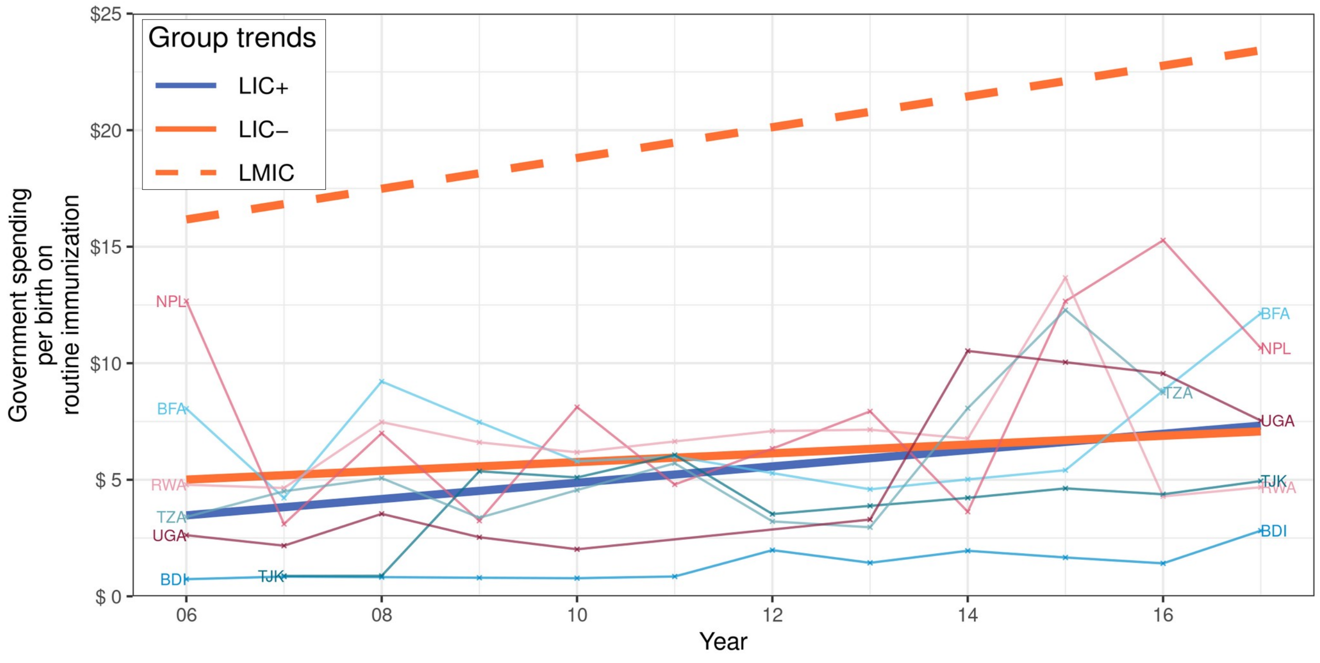


Figure 11: Total and government spending per birth on routine immunization of country groups – alternative country groups

Data source: IHME1. The trends of LIC+, LIC-, and LMIC were fitted by linear mixed-effects models. *LIC+ countries (ISO3)*: Burundi (BDI), Burkina Faso (BFA), Nepal (NPL), Rwanda (RWA), Tajikistan (TJK), Tanzania (TZA), and Uganda (UGA).

| Indicator (US$) | Year range | Starting trend value | | | Ending trend value | | | Yearly change rate | | |
|---|---|---|---|---|---|---|---|---|---|---|
| | | LIC+ | LIC- | LMIC | LIC+ | LIC- | LMIC | LIC+ | LIC- | LMIC |
| GNI per capita | 2000–18 | 184 | 320 | 571 | 841 | 802 | 2487 | 36·51 | 26·80 | 106·42 |
| GDP per capita | | 199 | 351 | 658 | 823 | 826 | 2576 | 34·66 | 26·40 | 106·57 |
| Total health spending per capita | 2000–16 | 22·67 | 31·72 | 62·75 | 45·68 | 42·64 | 110·55 | 1·44 | 0·68 | 2·99 |
| Government health spending per capita | | 5·79 | 9·77 | 23·51 | 11·33 | 7·15 | 47·98 | 0·35 | -0·16 | 1·53 |
| Private health spending per capita[✝] | | 12·92 | 19·43 | 35·55 | 18·70 | 22·37 | 53·96 | 0·36 | 0·18 | 1·15 |
| DAH per capita | | 4·02 | 2·60 | 3·71 | 15·56 | 13·12 | 8·42 | 0·72 | 0·66 | 0·29 |
| DAH per birth on newborn & child health | 2000–17 | 3·65 | 1·59 | 11·61 | 74·01 | 72·00 | 51·85 | 4·14 | 4·14 | 2·37 |
| DAH per birth on newborn & child health vaccines | | 5·57 | 1·64 | 2·25 | 28·14 | 28·31 | 18·93 | 1·33 | 1·57 | 0·98 |
| Total spending per birth on routine immunization | 2006–17 | 14·08 | 12·69 | 25·25 | 36·83 | 31·97 | 45·15 | 2·07 | 1·75 | 1·81 |
| Government spending per birth on routine immunization | | 3·47 | 5·01 | 16·17 | 7·32 | 7·08 | 23·43 | 0·35 | 0·19 | 0·66 |
| Total spending per birth on routine immunization vaccines | | 8·83 | 8·14 | 13·00 | 28·99 | 23·03 | 34·87 | 1·83 | 1·35 | 1·99 |
| Government spending per birth on routine immunization vaccines | | 2·07 | 4·26 | 9·87 | 4·19 | 4·12 | 19·46 | 0·19 | -0·01 | 0·87 |

*Table 4: Summary of financial trends of country groups – alternative country groups*

*Each indicator was fitted by a linear mixed-effects model. The table shows the values of the trends in the first and last years (intercepts at different times), and the yearly change rate over time (slope).*
*✝ Private health spending is the sum of out-of-pocket and prepaid private health spending.*

| **Indicator** | **LIC+ & LIC- comparison** | | | **LIC+ & LMIC comparison** | | |
|---|---|---|---|---|---|---|
| **(US$)** | **$\chi^2$** | **KR** | **PB** | **$\chi^2$** | **KR** | **PB** |
| GNI per capita | <0·0001* | 0·0003* | 0·0002* | <0·0001* | <0·0001* | <0·0001* |
| GDP per capita | 0·0013* | 0·0028* | 0·0014* | <0·0001* | <0·0001* | <0·0001* |
| Total health spending per capita | 0·0007* | 0·0017* | 0·0012* | <0·0001* | <0·0001* | <0·0001* |
| Government health spending per capita | <0·0001* | <0·0001* | <0·0001* | <0·0001* | <0·0001* | <0·0001* |
| Private health spending per capita[†] | 0·1666 | 0·1855 | 0·1839 | <0·0001* | <0·0001* | <0·0001* |
| DAH per capita | 0·6392 | 0·6599 | 0·6578 | <0·0001* | <0·0001* | <0·0001* |
| DAH per birth on newborn & child health | 0·9655 | 0·9688 | 0·9687 | 0·0007* | 0·0012* | 0·0008* |
| DAH per birth on newborn & child health vaccines | 0·3986 | 0·4177 | 0·4185 | 0·0152* | 0·0185* | 0·0158* |
| Total spending per birth on routine immunization | 0·5158 | 0·5412 | 0·5321 | 0·5555 | 0·5716 | 0·5707 |
| Government spending per birth on routine immunization | 0·4608 | 0·4730 | 0·4741 | 0·1551 | 0·1675 | 0·1628 |
| Total spending per birth on routine immunization vaccines | 0·2017 | 0·2236 | 0·2245 | 0·7680 | 0·7777 | 0·7725 |
| Government spending per birth on routine immunization vaccines | 0·1958 | 0·2117 | 0·2169 | 0·0177* | 0·0221* | 0·0205* |

*Table 5: Significance testing financial trends of LIC+ compared to LIC- and LMIC – alternative country groups*

*χ2, KR, and PB represent the p-values of an asymptotic $\chi^2$ test, a Kenward-Roger approximation for F tests for reduction of mean structure, and a parametric bootstrap method (10 000 simulations) respectively[6]. When a p-value is significant it means the group trends are significantly different to each other; i.e., it does not refer to the significance of a specific parameter of the regression but to the significance of all parameters combined.*

[†] *Private health spending is the sum of out-of-pocket and prepaid private health spending.*

* *p-values are significant with $p < 0{\cdot}05$.*
